# Symptom-specific genetics reveal heterogeneity within major depressive disorder

**DOI:** 10.64898/2026.03.24.26349158

**Authors:** Anastasia A. Goula, Floris Huider, Jouke-Jan Hottenga, Joëlle A. Pasman, Mariska Bot, M. Liset Rietman, Leen M. ’t Hart, Femke Rutters, Marieke T. Blom, Didi Rhebergen, Marjolein Visser, Catharina A. Hartman, Albertine J. Oldehinkel, Eco J. C. de Geus, Barbara Franke, H. Susan J. Picavet, W. M. Monique Verschuren, Hanna M. van Loo, Dorret I. Boomsma, Brenda W. J. H. Penninx, Yuri Milaneschi

## Abstract

**Background:** Major Depressive Disorder (MDD) is clinically and biologically heterogeneous. Here, we leveraged the genetics of individual depressive symptoms to dissect the disorder’s underlying heterogeneity.

**Methods:** We utilized the BIObanks Netherlands Internet Collaboration (BIONIC). A series of genome-wide association studies (effective-*N* range: 14,407 - 47,110) compared controls (N=48,286) with partially different subsets of lifetime MDD cases (range: 3,892–15,577), each endorsing one of 12 individual DSM-based depressive symptoms. Results were combined in genetic correlations that informed factor analyses with Genomic Structural Equation Modeling, decomposing underlying MDD liability dimensions. The identified factors were assessed and further characterized using multivariate regression of neurodevelopmental/psychiatric and cardiometabolic traits.

**Results:** All symptoms demonstrated substantial SNP-based heritability (*h²_SNP_:* 0.088 – 0.127). Despite high between-symptom genetic correlations, factor analyses yielded two highly correlated (*rg*=0.85) but still distinct latent factors: factor 1 (F1), capturing appetite/weight loss, insomnia, guilt/worthlessness, psychomotor slowing and suicidality, and factor 2 (F2), reflecting concentration problems, anhedonia, depressed mood, appetite/weight gain and fatigue. Overall, F1 had a stronger genetic overlap with neurodevelopmental/psychiatric phenotypes (e.g., autism: standardized estimate *β*=0.45, *p*=4.49×10⁻⁴; schizophrenia: *β*=0.40, *p*=1.73×10⁻⁴), while F2 significantly overlapped with cardiometabolic traits (e.g., metabolic syndrome: *β*=0.44, *p*=8.69×10⁻⁴; coronary artery disease: *β*=0.31, *p*=0.009).

**Conclusions:** We identified two genetic dimensions of MDD, each linked to partially distinct clinical manifestations and underlying biology, with one reflecting neurodevelopmental/psychiatric liabilities and the other capturing a strong cardiometabolic vulnerability. Disentangling such distinct dimensions may help guide patient stratification and targeted treatment, thereby advancing precision psychiatry.

## Introduction

Major Depressive Disorder (MDD) constitutes one of the leading causes of disability worldwide, with an estimated 12-month prevalence of 4–5% and prospective studies indicating a lifetime prevalence exceeding 30% (1). MDD is characterized by a complex multi-factorial etiology and is clinically heterogeneous, often showing comorbidity with other neurodevelopmental, psychiatric or cardiometabolic disorders (2). According to the Diagnostic and Statistical Manual of Mental Disorders, 5th Edition (DSM-5) (3), the range of depressive symptoms manifested by patients during MDD is diverse and may include opposing features, such as increases or decreases in appetite, weight, sleep, and psychomotor activity (4). It is conceivable that different clinical manifestations may represent the expression of distinct pathophysiological processes (5–7).

To dissect this heterogeneity, numerous efforts have focused on more homogeneous patient subgroups, also extending beyond symptoms to all possible biological and clinical aspects including biomarkers (8–10), genetic underpinnings (11,12), disease trajectories (13–15), treatment response (16,17), and comorbidities (18,19). The overarching goal is to identify robust, biologically informed depressive subtypes that will help advance patient stratification and targeted treatments in precision psychiatry (5,20). Nevertheless, no proposed subtyping framework has yet achieved broad consensus or consistent clinical application (21), highlighting the need for powerful methods and large datasets with detailed, clinically ascertained phenotyping.

From a genetic perspective, MDD is moderately heritable (*h*^2^=30%-40%) (22,23), with large-scale genome-wide association studies (GWAS) demonstrating its high polygenicity (24,25). Nevertheless, conventional GWAS treating MDD as a unitary construct may mask genetic signals that are specific to particular symptom dimensions (26). Indeed, prior research has indicated that individual depressive symptoms exhibit measurable SNP-based heritability and that their genetic architectures may partially diverge (27,28). This raises the possibility that the observed clinical heterogeneity in depression reflects, at least in part, underlying biological heterogeneity (26).

Recent advances in statistical genetics (26,29), such as Genomic Structural Equation Modeling (GenomicSEM) (30), now enable systematic exploration of this hypothesis by uncovering latent factors that may better capture the genetic architecture of MDD. However, progress has been limited due to the lack of large datasets with uniform, DSM-based granular phenotyping. To achieve adequate sample size, previous studies (31,32) examined the genetic bases of individual depressive symptoms without requiring a formal MDD diagnosis, potentially diluting the disorder-specific genetic signals. Additionally, some of those studies combined datasets including different diagnostic and self-report instruments (31), while others did not include granular symptom assessments, conflating symptoms with opposite directions into a single measure (32). As a result, identifying reproducible and biologically interpretable genetic dimensions of MDD has remained elusive.

Building on these limitations, the BIObanks Netherlands Internet Collaboration (BIONIC) (33) provides a uniquely well-suited resource. BIONIC combines a large, homogeneous sample with comprehensive, uniform DSM-based assessments of lifetime MDD and individual symptoms endorsed during the index major depressive episode, enabling analyses at a higher level of granularity and coherence. In this study, we leveraged BIONIC to perform GWAS of 12 DSM-based depressive symptoms endorsed during MDD, identify latent genetic factors, and characterize their external correlates with neurodevelopmental/psychiatric and cardiometabolic traits known to have a high affinity with the disorder. Our goal was to determine whether distinct genetic dimensions underlie specific symptom constellations and divergent pathophysiological pathways, thereby advancing precision psychiatry for MDD.

## Methods

### Study overview

Our study used a multi-step genomic approach, leveraging the genetic architecture of individual MDD symptoms (Figure 1). Upon conducting symptom-specific genome-wide-association studies (GWAS) in BIONIC, the summary statistics were annotated and analyzed for SNP-based heritability. Between-symptom genetic correlations were then derived to inform a GenomicSEM-based factor analysis modelling MDD latent dimensions. As a preliminary step for factor assessment, we examined the comparability of our data with those of previous GWAS studies examining individual depressive symptoms, followed by replication analyses. The BIONIC latent factors were further characterized through multivariate regression using GWAS of various neurodevelopmental/psychiatric and cardiometabolic traits. By regressing each external trait on all latent factors simultaneously, we estimated the unique contribution of each factor while accounting for their shared genetic influences. All analyses were performed using R (v4.4.2) (34) and Python (v3.11) (35).

**Figure 1.**
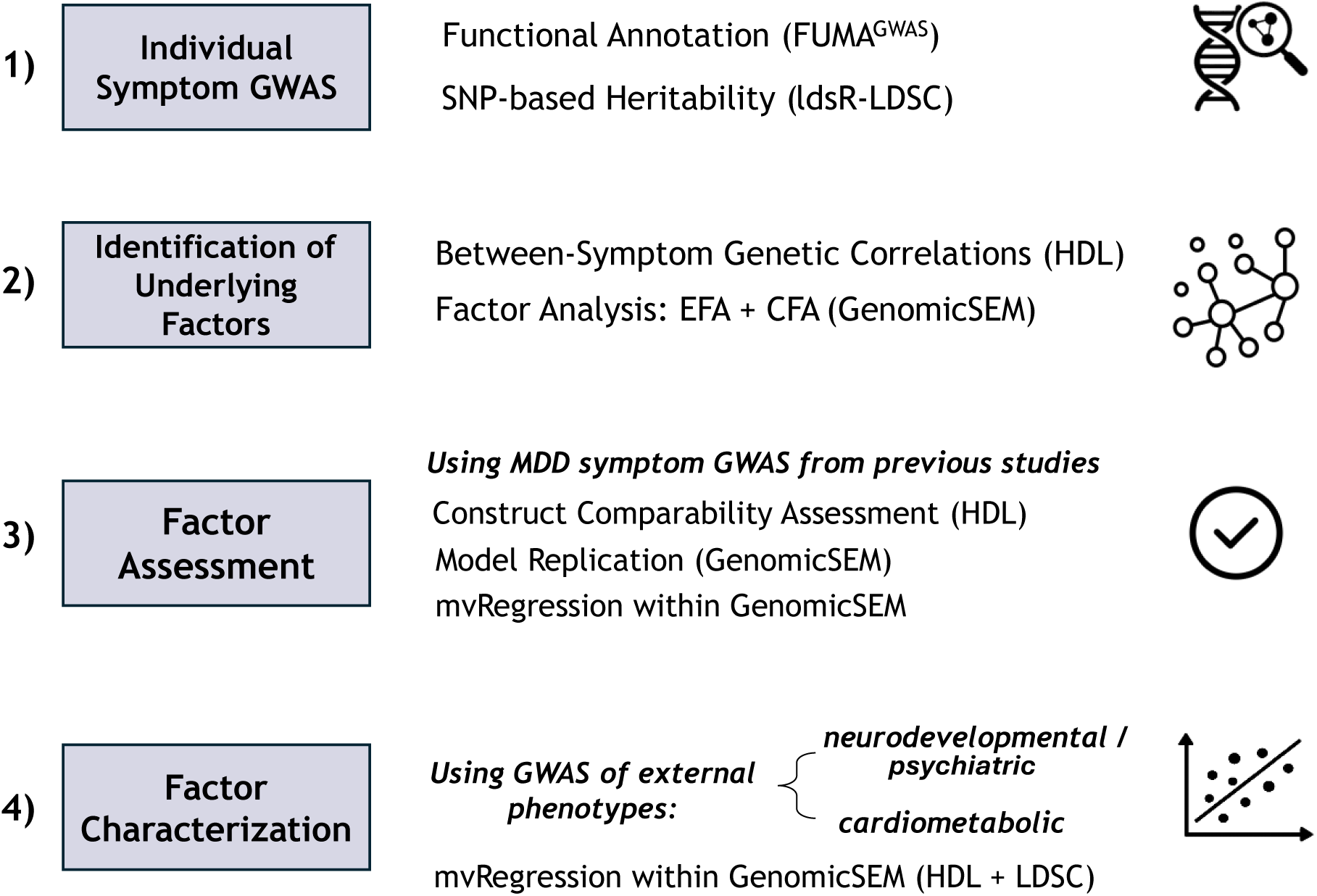
Schematic representation of our study and methodological approaches used. Abbreviations: GWAS: Genome-Wide Association Study, FUMA: Functional Mapping and Annotation, LDSC: Linkage Disequilibrium Score Regression, ldsR: R implementation of LDSC, HDL: High-Definition Likelihood, EFA: Exploratory Factor Analysis, CFA: Confirmatory Factor Analysis, GenomicSEM: Genomic Structural Equation Modeling, MDD: Major Depressive Disorder

### Study sample: BIONIC

The study sample was derived from the BIObanks Netherlands Internet Collaboration (BIONIC) project, a nation-wide initiative involving 16 Dutch cohorts, aimed at achieving uniform and in-depth phenotyping for MDD (33). A detailed description of the project rationale and methods, including harmonization of phenotypic and genetic data, has been previously given in the full cohort paper (33) and the main BIONIC MDD GWAS mega-analysis (36).

The starting sample for the present analyses is the same that was used in the main BIONIC MDD GWAS (36): 16,655 participants with lifetime MDD (established according to DSM-5 criteria) and 48,286 screened controls, free of MDD and any major psychiatric disorders. In the present study, we leveraged additional information available for MDD cases, i.e., the endorsement during the index major depressive episode of twelve symptoms mapping to DSM A1-A9 criteria for MDD. This additional data was fully available only in MDD cases due to the structured skip pattern in the assessment tools, enabling the evaluation of the whole set of DSM symptoms only in participants endorsing at least one among the core A1/A2 MDD criteria. Assessed symptoms included the following: depressed mood (DepMood), diminished interest or pleasure (Anhedonia), appetite or weight loss (AppWLoss), appetite or weight gain (AppWGain), diminished sleep (Insomnia), excessive sleep (Hypersomnia), psychomotor slowing (Slowing), psychomotor agitation (Agitation), energy loss (Fatigue), feelings of worthlessness or guilt (Guilt/Worth), concentration problems or indecisiveness (ConcProb), and thoughts about death or suicide (Suicidality). Among the 16,655 lifetime MDD cases, the number of subjects endorsing each of the 12 symptoms varied from 3,892 (Slowing) to 15,577 (Fatigue). Full prevalences are reported in Table 1.

**Table 1.** Baseline characteristics of BIONIC participants for each symptom-MDD GWAS. Cases are defined as individuals with MDD also endorsing a specific depressive symptom, while controls are participants without MDD, irrespective of specific symptom endorsement (36). *N_total_* = *N_controls_* (48,286) + *N_cases_* (range: 3,892 - 15,577, depending on the symptom in question); *N_MDD_* = 16,655; *N_eff_* = 4 / [(1/*N_cases_*) + (1/*N_controls_*)] (40)

| Symptom | Abbreviations | Symptom<br>Prevalence<br>in MDD<br>$N_{cases}/N_{MDD}$ | $N_{eff}$ | $N_{cases}$<br>(% of $N_{total}$) | Mean Age<br>(SD) | Females<br>(%) |
| --- | --- | --- | --- | --- | --- | --- |
| 1. Depressed Mood | DepMood | 0.908 | 46072 | 15126 (23.9) | 50.8 (16.0) | 60.6 |
| 2. Diminished Interest or Pleasure | Anhedonia | 0.909 | 46120 | 15147 (23.9) | 50.8 (16.0) | 60.5 |
| 3a. Appetite Decrease or Weight Loss | AppWLoss | 0.443 | 25615 | 7383 (13.3) | 51.2 (16.3) | 59.5 |
| 3b. Appetite Increase or Weight Gain | AppWGain | 0.243 | 14943 | 4049 (7.7) | 51.1 (16.3) | 59.0 |
| 4a. Diminished Sleep (Insomnia) | Insomnia | 0.503 | 28540 | 8372 (14.8) | 51.2 (16.2) | 59.4 |
| 4b. Excessive Sleep (Hypersomnia) | Hypersomnia | 0.281 | 17073 | 4682 (8.8) | 50.9 (16.4) | 58.6 |
| 5a. Psychomotor Slowing | Slowing | 0.234 | 14407 | 3892 (7.5) | 51.3 (16.3) | 58.4 |
| 5b. Psychomotor Agitation | Agitation | 0.270 | 16439 | 4492 (8.5) | 51.3 (16.3) | 58.5 |
| 6. Energy Loss or Fatigue | Fatigue | 0.935 | 47110 | 15577 (24.4) | 50.7 (16.0) | 60.8 |
| 7. Feelings of Worthlessness or Guilt | Guilt/Worth | 0.750 | 39705 | 12495 (20.6) | 50.7 (16.1) | 60.4 |
| 8. Concentration Problems or Indecisiveness | ConcProb | 0.922 | 46601 | 15355 (24.1) | 50.8 (16.0) | 60.5 |
| 9. Recurrent Thoughts of Death or Suicide (Suicidality) | Suicidality | 0.498 | 28307 | 8292 (14.7) | 51.1 (16.2) | 59.4 |

### Individual-symptom GWAS

We performed a series of twelve GWAS comparing the same group of controls with twelve partially different subsets of lifetime MDD cases. For each subset, we selected MDD cases endorsing one of the 12 individual symptoms during the index major depressive episode (Supplemental Figure 1). Analyses were performed using *fastGWA* (37) from the *GCTA* software (38) with generalized linear mixed models, correcting for sex, age, 10 ancestry-informative principal components, genotyping array, and relatedness (via a genetic relationship matrix).

The resulting GWAS summary statistics underwent standard quality control procedures and annotation (S1). SNP-based heritability (*h*^2^_SNP_) was estimated using *ldsR* (https://github.com/Ararder/ldsR) (39), an R implementation of LDSC (40). To facilitate comparability across symptoms with differing prevalences, the *h*^2^_SNP_ was derived on the observed scale assuming a 50/50 case-control distribution as done previously (41). For reference, we used the HapMap3 variants of the European sample of 1000 Genomes Phase 3 (42).

SNP-based genetic correlations between symptoms were estimated using High-Definition Likelihood (HDL; https://github.com/zhenin/HDL/) (43), an approach well-suited for analyzing summary statistics from smaller-scale GWAS (44). *P*-values for all correlations were adjusted using the false discovery rate (FDR) method.

### Identification of underlying factors

Our factor analysis consisted of an exploratory and a confirmatory stage. To derive genetically informed factors, we fit structural equation models using the *GenomicSEM* (30) package in R (34). After harmonizing the summary statistics using the default parameters and HapMap3 as the reference, we recalculated the HDL genetic correlations within GenomicSEM. To this end, we rescaled the symptom prevalences observed among MDD cases in the BIONIC dataset by the estimated population prevalence of lifetime MDD in the Netherlands (∼25%) (45). For the exploratory factor analysis (EFA), we examined models with one to five factors (*n*=1–5) using the *factanal* function in R. In the confirmatory factor analysis (CFA), we applied a loading threshold of 0.3, allowed cross-loadings, and used standard fit indices to evaluate each model. Those include Comparative Fit Index (*CFI*), Standardized Root Mean Square Residual (*SRMR*), Akaike Information Criterion (*AIC*), and the chi-square statistic (*χ²*). To inform factor retention, we conducted parallel analysis using the *paLDSC* function of the *GenomicSEM* package by simulating random genetic correlation matrices matched to the observed data and comparing their eigenvalues to those of the empirical matrix. Model selection was based on convergence of evidence from parallel analysis and global fit indices, prioritizing solutions that achieved adequate fit while minimizing unnecessary complexity. More information on the factor analysis can be found in S2.

### Factor Assessment

To assess the identified factors, we leveraged two previous independent studies that also performed GWAS of individual MDD symptoms. The first was a meta-analysis (32) primarily based on UK Biobank (UKB) (46), also including data from the GLAD (47) and PROTECT (48) cohorts (UKB_meta, *N* = 224,535 – 308,421). In contrast to BIONIC, this study conducted GWAS of individual depressive symptoms as continuous traits, rather than examining them within the context of an MDD diagnosis. Additionally, as phenotyping was based on the PHQ-9, opposite symptoms were conflated in single items examining generic change during a depressive episode (e.g., Appetite/Weight Changes, Sleep Problems, Psychomotor Changes). Secondly, summary statistics for individual MDD symptoms were also derived by the Psychiatric Genomics Consortium (PGC), which used similarly disaggregated symptoms as BIONIC (e.g. distinguishing Insomnia from Hypersomnia). However, different case-control definitions were used: cases were defined based solely on symptom endorsement, without requiring an MDD diagnosis. Similarly, controls were defined as participants not endorsing the symptom in question, without being screened for MDD. It is worth noting that the PGC conducted separate analyses in MDD case-enriched (PGC_ca, *N_eff_* =2,471 - 11,130) and community-based (PGC_com, *N_eff_* =218 - 102,071) cohorts.

As a preliminary step, we evaluated the comparability of the BIONIC constructs with those of the other available datasets (UKB_meta, PGC_ca, PGC_com) by computing HDL genetic correlations between similar symptoms assessed across studies. Furthermore, by using GenomicSEM, the model identified in BIONIC was fitted in both PGC_ca and PGC_com. Given the challenges of replicating factor structures across datasets (due to varying case definitions, symptom measures, and the inherent constraints of Genomic SEM), we additionally performed genetic multivariate regression analyses (mvReg) as an alternative indirect assessment of the BIONIC-derived factors. To this end, we mainly utilized the well-powered UKB_meta data and, for the disaggregated symptoms, we opted for the MDD-enriched PGC_ca data, aiming to approximate BIONIC’s case definition (i.e., symptom endorsement together with MDD). These external GWAS summary statistics for depressive phenotypes (UKB_meta and PGC_ca) were regressed onto both BIONIC-derived latent factors simultaneously using genetic multivariate regression (External Trait ∼ F1 F2), enabling assessment of their relative genetic associations while accounting for shared variance. We report standardized estimates (predictor-standardized), reflecting the effect of a 1-*SD* increase in each genetic factor on the raw symptom scale. More information on the external studies and factor assessment can be found in S3.

### Factor Characterization

To elucidate the underlying biology of each factor, we also performed mvReg on both BIONIC-derived factors with various external phenotypes from two major classes: “neurodevelopmental/psychiatric disorders” (Attention-Deficit/Hyperactivity Disorder [ADHD], Autism Spectrum Disorder [ASD], Bipolar Disorder [BIP], Major Depressive Disorder [MDD], Schizophrenia [SCZ]), and “cardiometabolic traits” (Body Mass Index [BMI], Coronary Artery Disease [CAD], C-Reactive Protein [CRP], Type-2 diabetes [DiabetesII], Glycoprotein Acetyls [GlycAcetyl], Glycated Hemoglobin [HbA1c], Interleukin-6 [IL-6], Leptin, Metabolic Syndrome [MetSyndr], Stroke, Triglycerides [Triglyc]). These traits were chosen as representative, well-characterized examples with highly-powered GWAS data. More information on the summary statistics for each trait can be found in S4.1 (Supplemental Tables 5 and 6).

Genetic correlations were first estimated with HDL, as in earlier steps. Since for many traits the *rg* was not estimable with that method, we additionally employed LDSC as a complementary approach (see S4.2). The output of each method was then used to regress the external traits on both factors simultaneously, yielding the external correlates of each factor while accounting for the presence of the other (as in Factor Assessment).

## Results

### Individual-symptom GWAS

The main characteristics of our study sample per individual MDD symptom are summarized in Table 1. Across all traits, the mean effective sample size (*N_eff_*) was 30,911 (range: 14,407 – 47,110), with a mean age of 50.97 years and a higher proportion of females (59.74%). Genomic inflation factors (*λ_GC_*) of the GWAS ranged from 1.035–1.091 (*λ_1000_*=1.004–1.006) and LDSC Intercepts were near 1 (0.992 - 1.016), indicating the absence of population stratification (ST1). Except for two loci (*rs3818852* for DepMood and *rs147067514* for Agitation, see S1 and ST2), most GWAS did not yield genome-wide significant SNPs. This was expected given the limited sample size and therefore reduced statistical power. Importantly, the identification of genome-wide significant variants was not the primary objective of this study, rather than characterizing the overall polygenic architecture of each MDD symptom. Moreover, we note that contrasting symptom-level GWAS against the same control group may inflate significance estimates, warranting cautious interpretation of individual SNP findings (49).

Each individual-symptom GWAS captured considerable SNP heritability (*h^2^_SNP_*, see diagonal of Figure 2). Across all depressive symptoms, the most heritable were suicidality (0.127), followed by appetite/weight gain (0.124) and psychomotor agitation (0.115), while the least heritable were depressed mood (0.090) and fatigue (0.090). Most *h²_SNP_ Z*-scores were >4, with four symptoms showing lower values (AppWGain, *Z*=3.81; Hypersomnia, *Z*=3.59; Agitation, *Z*=3.40; and Slowing, *Z*=2.93). Nevertheless, three of these were still above *Z*=3, indicating that the respective estimates, although less powered, remained meaningfully interpretable.

**Figure 2.**
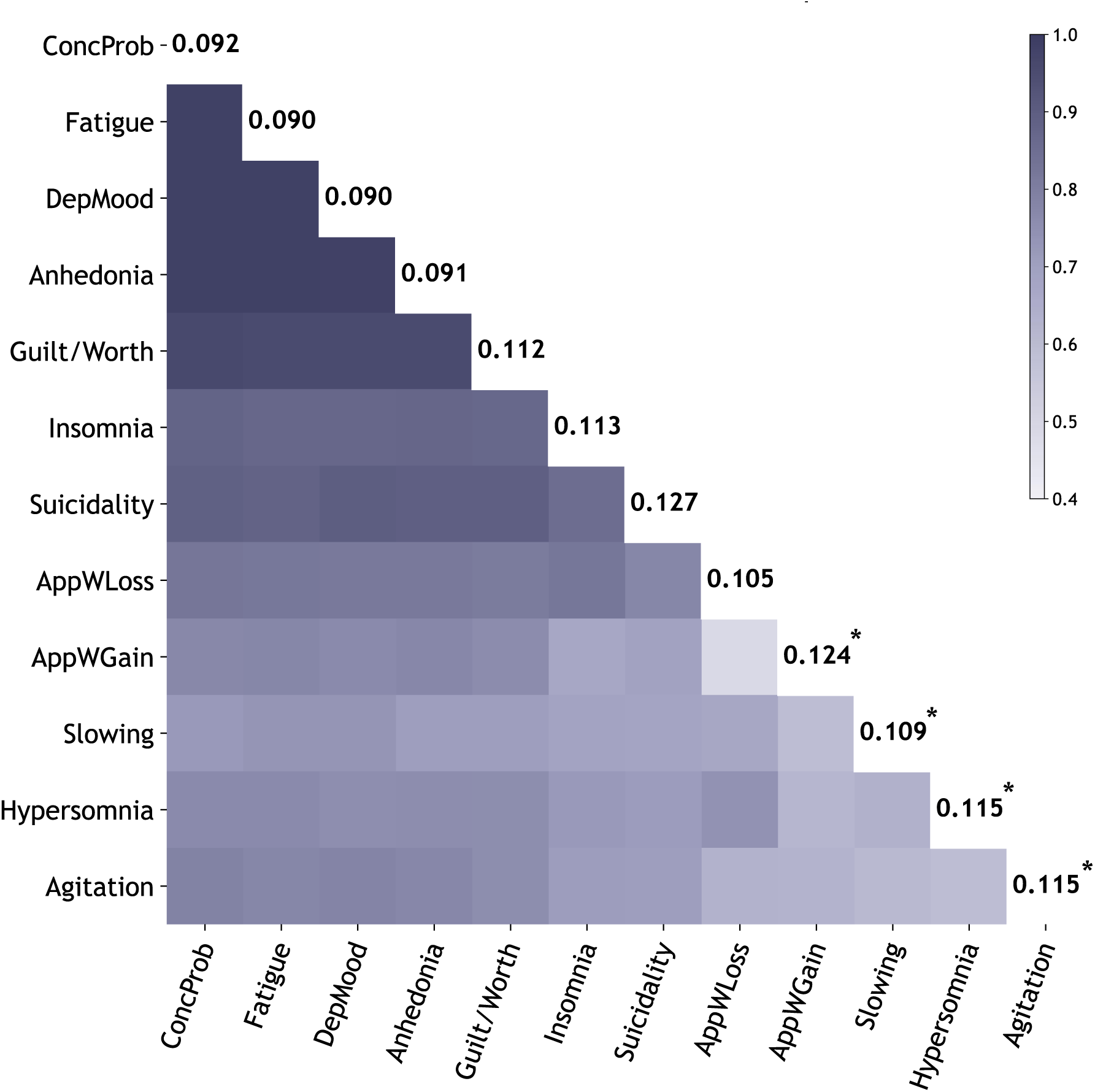
Hierarchically clustered HDL genetic correlations between depressive symptoms within an MDD diagnosis. Correlation estimates ranged from 0.49 to 0.98 and were all significant (*p_fdr_*<0.05). The numbers on the diagonal represent the estimated observed-scale SNP heritability of each individual symptom within MDD. Symptoms with *Z*-scores <4 are marked with asterisks(*).

### Identification of underlying factors

The HDL correlations were consistently high and positive (Figure 2, ST3). This was expected considering they were based on twelve GWAS that compared the same group of controls with partially overlapping subsets of lifetime MDD cases endorsing specific symptoms. Despite this overarching homogeneity in case-control contrast across GWAS, relatively lower genetic correlations were found between specific symptom pairs. The lowest correlation estimate was observed between MDD with AppWLoss and MDD with AppWGain (*rg*=0.49, *se*=0.03), which was statistically different from 1 (*p_1_*=3.94×10^-52^). This suggests that, within MDD cases, those manifesting AppWGain versus AppWLoss during the index depressive episode were the most genetically distinct subgroups.

Parallel analysis indicated that the optimal number of factors was identified prior to the two-factor solution (Supplemental Figure 3), consistent with a dominant general genetic factor underlying depressive symptoms within MDD. However, subsequent factor analyses demonstrated that a two-factor solution improved model fit and enabled meaningful differentiation between symptom dimensions (S.2.7). Model selection was guided by the fit–parsimony trade-off, aiming to balance adequate model performance with simplicity. Thus, while acknowledging a strong general liability, we retained the two-factor model for subsequent analyses to preserve both structural nuance and interpretability. This allowed for greater discrimination and the potential identification of more specific genetic components within MDD.

In CFA, the EFA-derived two-factor model showed moderate to excellent fit indices (*CFI* = 0.89, *SRMR* = 0.04, *AIC*= 5455366, *Chisq*= 5455312) and comprised two highly correlated latent factors (*rg*=0.86, *p_1_*=9×10^-4^) (Figure 3). One factor (F1) showed statistically significant loadings of insomnia, feelings of worthlessness/guilt, appetite/weight loss, suicidality, psychomotor slowing and hypersomnia. The second factor (F2) had significant loadings of concentration problems, anhedonia, depressed mood, appetite/weight gain and fatigue. Psychomotor agitation exhibited no significant loadings (Figure 3). More information on the structure and performance metrics of all tested models can be found in S2.7 and ST5a-e.

**Figure 3.**
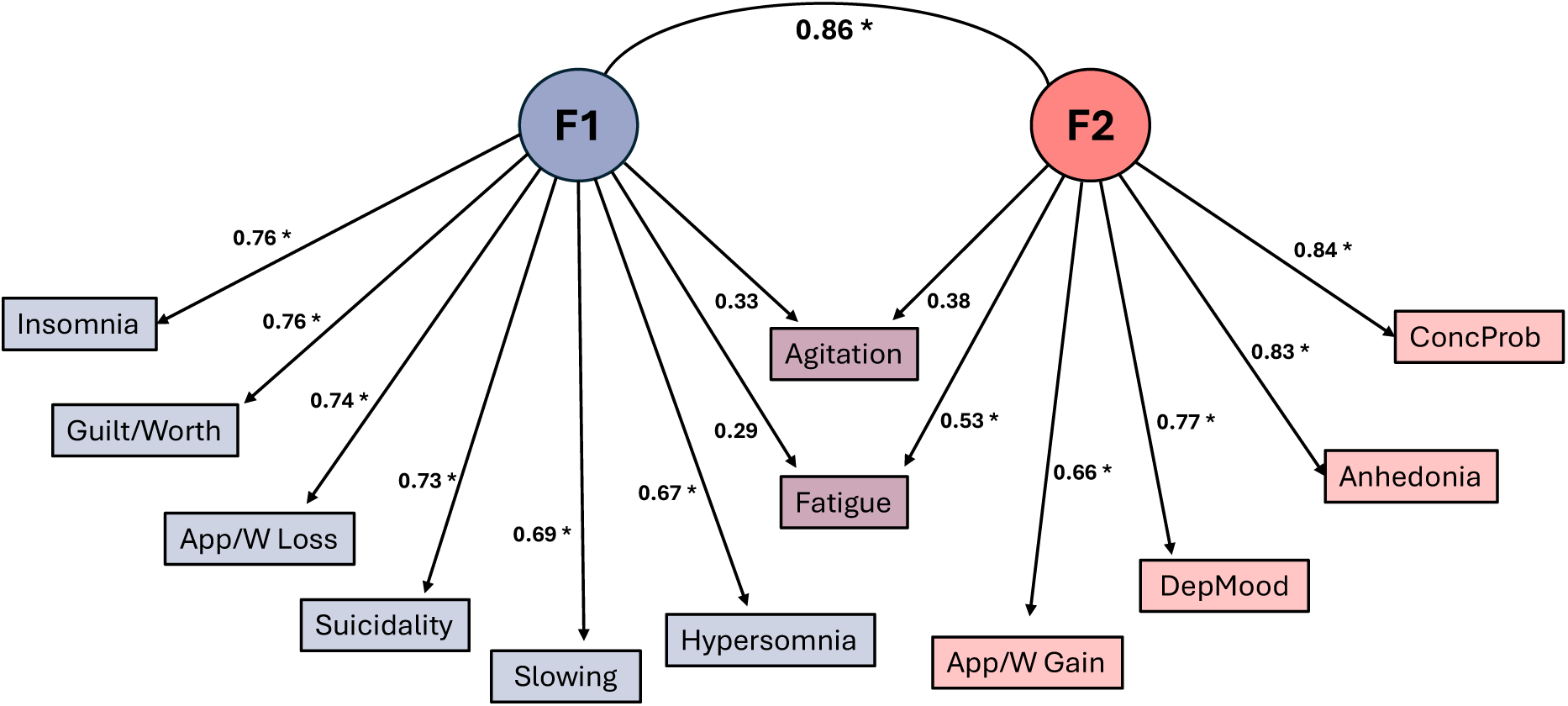
The two-factor model that was selected from the factor analysis. Significant (p<0.05) results are marked by asterisks (*).

### Factor Assessment

We evaluated the comparability of the BIONIC data with those of previous studies by estimating genetic correlations with similar individual depressive symptoms assessed in UKB_meta and PGC (S3.2). For UKB_meta, among the estimable same-symptom genetic correlations, estimates ranged from 0.62 (Fatigue) to 0.77 (Suicidality). In the case-enriched dataset from the PGC, estimable correlations ranged from 0.22 (AppWLoss, Insomnia) to 0.95 (Guilt/Worth), while in the community-based PGC dataset correlations ranged from 0.43 (AppWLoss) to 0.77 (Hypersomnia). Overall, while statistically significant findings were observed for most same-symptom genetic correlations, the presence of low estimates indicated a substantially limited comparability between BIONC data and those of other studies. This was likely due to inherent methodological differences including differing GWAS designs, case-control definitions and symptom measurement instruments. The lack of data compatibility across studies substantially limited the possibility of validation for the BONIC model using the previous GWAS.

Replication of the factor model via CFA was feasible in both PGC_ca and PGC_com, which, similarly to BIONIC, are characterized by finer-grained, directional symptom definitions. Nevertheless, both models’ fit was suboptimal, with poor fit indices (case-enriched: *CFI*=0.456, *SRMR*=0.220; community-based: *CFI*=0.777, *SRMR*=0.136), highlighting challenges in reproducing exact factor structures across studies with differing case definitions and cohort compositions (S3.4).

As an alternative, indirect assessment, we next used mvReg within GenomicSEM to explore patterns of genetic associations between the external symptoms and the factors identified in BIONIC. Consistently with the original model, depressed mood, concentration problems and fatigue were significantly associated with Factor 2, while suicidality linked to Factor 1. Although not statistically significant, likely due to the small samples of the PGC GWAS, the two most genetically divergent symptoms of appetite/weight loss and gain (Figure 3) showed suggestive divergent associations with the two BIONIC factors: appetite/weight loss positively related to F1 and negatively to F2, while appetite/weight gain was positively associated with F2 and negatively with F1 (Figure 4, ST7).

**Figure 4.**
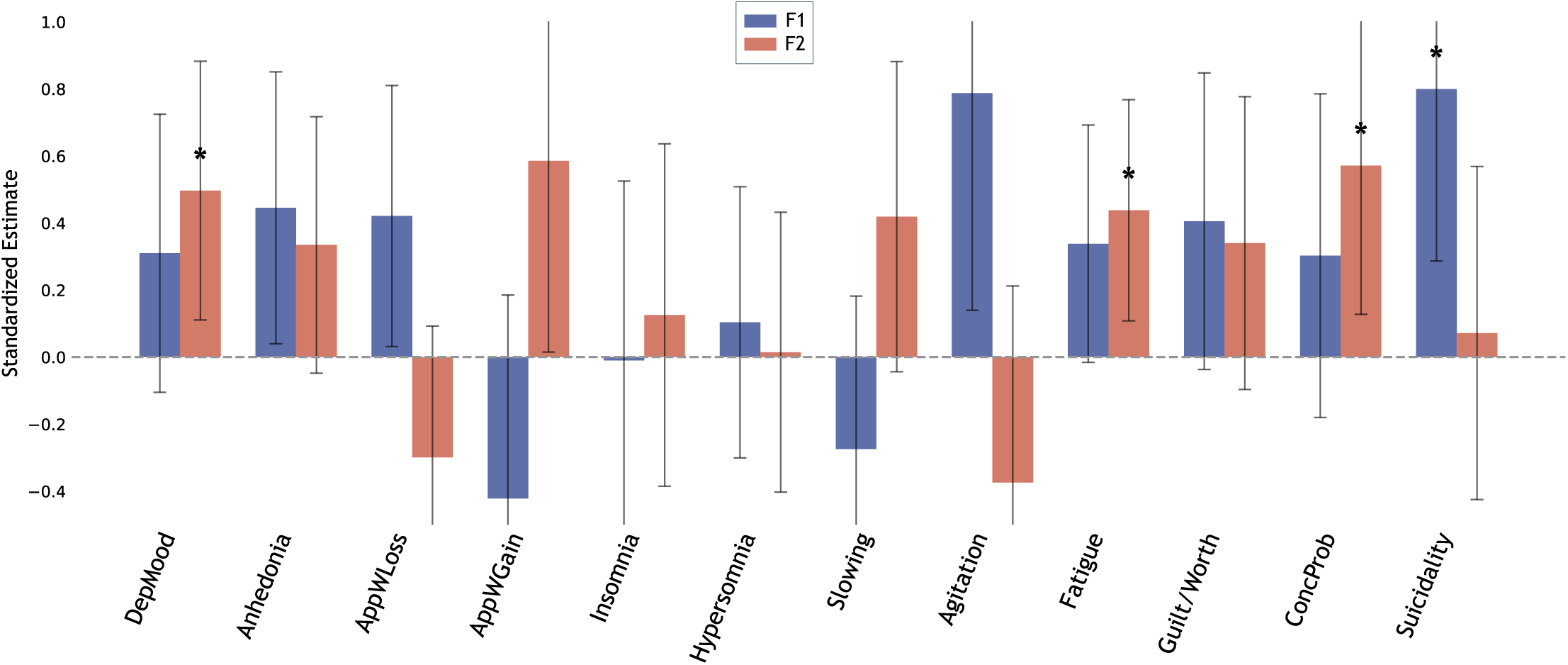
Multivariate regression of external GWAS genetic signals for MDD symptoms on the latent factors derived by BIONIC. Error bars represent 95% confidence intervals around the standardized effect (predictor standardized; outcome unstandardized). Significant results (*p*<0.05) are indicated with asterisks (*).

### Factor Characterization

To further characterize the two latent factors, genetic liabilities for external traits across neurodevelopmental/psychiatric and cardiometabolic domains were regressed on both factors simultaneously within the GenomicSEM framework (ST8). In analyses based on genetic correlation matrices derived from HDL (Figure 5, upper panel), neurodevelopmental/psychiatric disorders showed stronger and statistically significant associations with F1 (e.g., autism: *β*=0.45, *p*=4.49×10⁻⁴; bipolar disorder: *β*=0.44, *p*=7.72×10⁻^5^), with ADHD being the sole exception demonstrating the opposite pattern with a significant association with F2 (*β*=0.49, *p*=0.002). In contrast, cardiometabolic traits tended to associate more strongly with F2, with several significant positive effects for CAD (*β*=0.31, *p*=0.009), CRP (*β*=0.32, *p*=0.013), leptin, (*β*=1.14, *p*=0.041) and metabolic syndrome (*β*=0.44, *p*=8.69×10⁻⁴;), which was also negatively associated with F1 (*β*=-0.28, *p*=0.049). Except for leptin, all other associations remained significant after correcting for multiple testing (ST8a). Since several traits (BMI, DiabetesII, interleukin-6, triglycerides) could not be fitted with HDL-based matrices, potentially reflecting collinearity-related limitations of the latter method (S4.2), we repeated the analyses using matrices based on the LDSC method (Figure 5, lower panel). The profile of the associations was consistent with those based on HDL, although with larger variance intervals as expected due to the lower precision of LDSC. BMI showed significant and divergent associations with the factors, negative with F1 (*β*=-1.70, *p*= 0.042) and positive with F2 (*β*=1.75, *p*= 0.036); triglyceride levels were positively associated with F2 (*β*=1.03, *p*=0.046). None of these associations remained significant after correcting for multiple testing (ST8b).

**Figure 5.**
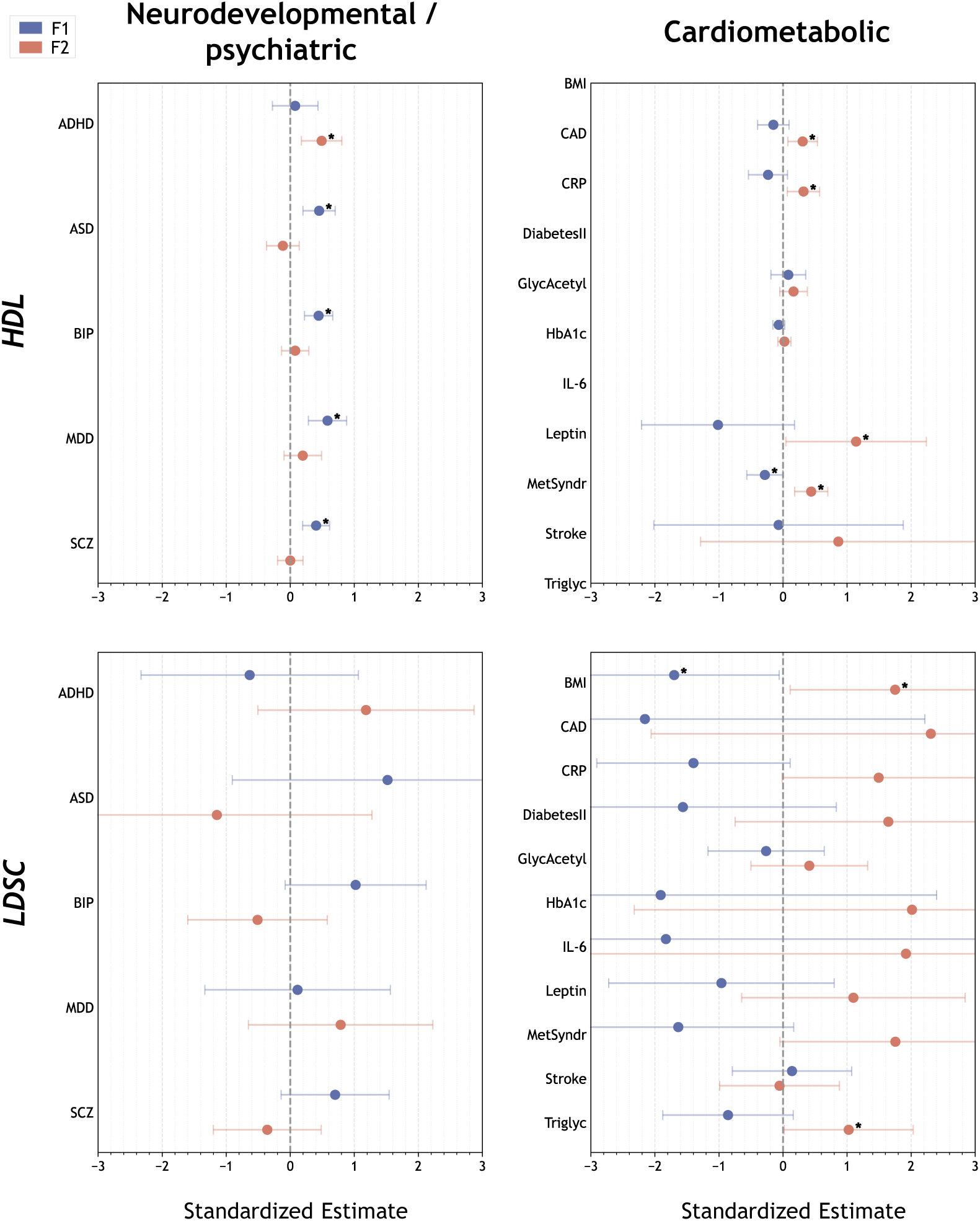
Multivariate regression of external neurodevelopmental/psychiatric (left) and cardiometabolic (right) phenotypes on the GenomicSEM-derived latent factors (F1: blue, F2: red) using HDL (top) and LDSC (bottom). Error bars represent 95% confidence intervals around the standardized effect (predictor standardized; outcome unstandardized). Significant results (*p*<0.05) are indicated with asterisks (*). External phenotypes: Attention-Deficit/Hyperactivity Disorder (ADHD), Autism Spectrum Disorder (ASD), Bipolar Disorder (BIP), Major Depressive Disorder (MDD), Schizophrenia (SCZ), Body Mass Index (BMI), Coronary Artery Disease (CAD), C-Reactive Protein (CRP), Type-2 diabetes (DiabetesII), Glycoprotein Acetyls (GlycAcetyl), Glycated Hemoglobin (HbA1c), Interleukin 6 (IL-6), Leptin, Metabolic Syndrome (MetSyndr), Stroke, Triglycerides (Triglyc)

## Discussion

In this study, we leveraged the Dutch BIONIC cohort to dissect MDD heterogeneity. BIONIC comprises ∼48,500 screened controls and >15,000 individuals with lifetime MDD, diagnosed through a uniform DSM-based assessment and complemented by a granular evaluation of symptoms manifested during the index depressive episode. By performing a data-driven analysis utilizing the genetic signature of individual, DSM-based depressive symptoms, we identified two partially distinct genetic dimensions underlying MDD; each dimension was linked to the expression of a specific clinical profile and showed highly divergent patterns of genetic overlap with neurodevelopmental/psychiatric and cardiometabolic traits. Overall, our findings suggest the existence of partially divergent genetic dimensions underlying MDD, highlighting the disorder’s heterogeneity and inherent complexity.

All symptom GWAS captured substantial heritability (*h²_SNP_* ≈0.09–0.13), demonstrating that the expression of each symptom in the context of MDD is partially driven by genetic factors. Building on the symptom-level genetic signal, genetic correlation analyses showed a substantial coherence of the MDD diagnosis, with high genetic overlap (median *rg*=0.80) between individual symptoms and factor analyses pointing to a unitary construct that could be reliably decomposed only in two underlying dimensions. This substantial genetic coherence of DSM-based MDD is in contrast with claims related to the lack of consistency of the disorder (50). Nevertheless, we also found noticeable genetic divergence within MDD; for instance, cases manifesting appetite increase during the index depressive episode were the least genetically similar to cases manifesting appetite decrease (*rg*=0.49, significantly different from 1). This is consistent with previous studies showing that the direction of appetite alterations during depression reliably discriminates patient groups with differing genetic, biological and neural profiles (12,51–55).

To further examine how these patterns relate to the broader question of MDD dimensions and their underlying structures, genomic factor analysis was applied. We identified two partially distinct, but highly correlated latent factors. The first factor (F1) predominantly reflected melancholic or anxious depression traits, including appetite/weight loss, insomnia, guilt/worthlessness, hypersomnia, psychomotor slowing, and suicidality. The second factor (F2) was largely characterized by atypical, cognitive and motivational symptoms, including concentration problems, anhedonia, depressed mood, appetite/weight gain and fatigue.

In this genetically-informed study, our findings contribute to the growing, though still inconsistent, body of (genetic) research aimed at disentangling depression’s heterogeneity (20,31,56,57). Much of this inconsistency may stem from variation in study design, diagnostic assessments, and sample characteristics across studies. These sources of variation shape both the clinical expression of depression and the genetic architecture captured in each cohort, thereby hindering the reproducibility of symptom-level factors or genetic dimensions across studies. Consistent with this challenge, the replication of our factors was inherently constrained, particularly given the differences in case definitions and the level of phenotypic granularity in currently available GWAS.

To further characterize the genetic signatures of the identified factors, we performed multivariate regression analyses incorporating a range of external neurodevelopmental/psychiatric and cardiometabolic traits known to be linked to MDD. Overall, F1 was more strongly associated with neurodevelopmental/psychiatric traits such as schizophrenia or autism spectrum disorder, whereas F2 showed stronger links to cardiometabolic phenotypes such as BMI or triglycerides. Interestingly, among the neurodevelopmental/psychiatric disorders, Attention Deficit/Hyperactivity Disorder (ADHD) was the only phenotype more closely related with F2 rather than F1. This aligns with previous genetic studies suggesting that both ADHD and MDD are characterized by substantial hormonal and metabolic influences, primarily involving insulin-related pathways in the brain (58,59). Regarding cardiometabolic traits, F2 was positively and significantly associated with BMI and metabolic syndrome, whereas F1 showed negative associations with these traits. Additional unique positive associations for F2 included the inflammatory markers CRP and IL-6, triglycerides, type II diabetes and the hormone leptin, all reinforcing the factor’s connection to metabolic and inflammatory processes, as indicated previously (12,60,61). Finally, the association of CAD with F2 but not F1 supports prior evidence of an MDD subtype having stronger associations with adverse cardiometabolic profiles/outcomes compared to overall MDD (62,63).

These findings add further support to the model of “immunometabolic depression”, which has been gaining increasing attention in research (4,64). In line with previous studies (11,65), our results point to a biologically coherent MDD dimension (F2) characterized predominantly by motivational and energy-related symptoms (anhedonia, fatigue, increased appetite or weight gain, concentration problems) and intertwined inflammatory, cardiometabolic, and energetic dysregulation. Such findings have been reported across multiple methodological domains, including omics signatures (61–63,66–68), clinical profiles (69–71), neuroimaging (72,73), and even integrative multi-domain approaches (74), which are emerging as promising research directions (75,76).

Our study has several strengths. First, all data were sourced from BIONIC (33), a nationwide initiative that applies uniform and validated phenotyping in a homogenous population. By ensuring standardized and reliable case assessment, BIONIC provides a solid foundation for our analyses. Additionally, by capturing the specific direction of symptom change during MDD (e.g. Appetite/Weight Loss vs Gain), BIONIC provides the resolution needed to detect differential genetic signals or mechanisms that would otherwise be lost in aggregated measures (6,31,77). As for our analyses, an important strength is that we allowed the symptom composition of each latent factor to emerge freely from the genetic correlation patterns. Despite not constraining the models a priori, this approach still yielded interpretable and biologically meaningful latent factors converging with findings and theoretical developments in the field (4,65).

Our study also has some limitations. First, since BIONIC’s original purpose was to examine a well-characterized, DSM-defined MDD phenotype in a homogenous Dutch European population, extrapolation of the present findings to populations of different ancestral backgrounds is not possible. Additionally, while GenomicSEM provides a powerful framework for capturing latent genetic structures, there are also some inherent technical caveats: susceptibility to overfitting and factor indeterminacy (30,78). In light of these limitations, we conducted a study without restricting the models a priori and allowing symptoms to load freely on the latent factors. Finally, due to major methodological and conceptual differences between our data and those available from previous GWAS, we had limited possibility to validate and confirm the main findings in independent datasets. Thus, replication of the suggested model in data from independent, similarly characterized cohorts (i.e., comparable case-control definitions and data structures) will be critical to fully substantiate our findings and assess their applicability in broad clinical practice.

In summary, our findings highlight the multidimensional genetic architecture of MDD, investigating two latent factors with distinct symptom profiles and biological mechanisms. These findings not only support prior hypotheses about distinct underlying bio-clinical dimensions of MDD but also uncover previously unrecognized patterns of genetic differentiation across symptom domains, their links to systemic biology, and their potential relevance for comorbid psychiatric and somatic conditions. Importantly, the identification of a cardiometabolic-related dimension reinforces the concept of MDD as not solely a brain disorder, but a systemic condition with links to metabolic and inflammatory processes. By dissecting symptom heterogeneity and their genetic underpinnings, this study lays the groundwork for future precision medicine approaches aimed at more targeted prevention, stratification, and treatment. Validation in diverse, fully genotyped cohorts integrating genomic, phenotypic, and other biological data will be essential to translate these findings into clinically actionable strategies.

## Data Availability

Summary statistics and analysis code will be made publicly available upon publication.

## Acknowledgements

YM is partially supported by Amsterdam UMC StarterGrant (Ronde2), Amsterdam Neuroscience (PoC funding 2024-2026), and the Immuno MIND consortium, funded by UK Research and Innovation as part of the UK national Mental Health Platform. HMvL was supported in part by a VENI grant from the Talent Program of the Netherlands Organization of Scientific Research (NWO-ZonMW 09150161810021) and by NIMH grant R01MH125902.

We are very grateful to everyone who participated in this research or worked on this project and its contributing studies. Funding for the BIONIC project was awarded to Dorret Boomsma and Brenda Penninx by the Biobanking and Biomolecular Resources Research Infrastructure (BBMRI-NL: 184.021.007; 184.033.111). Below are cohort-specific funding declarations and acknowledgements. We would like to thank the research participants and employees of 23andMe for making this work possible. Lifelines The Lifelines initiative has been made possible by subsidy from the Dutch Ministry of Health, Welfare and Sport, the Dutch Ministry of Economic Affairs, the University Medical Center Groningen (UMCG), Groningen University and the Provinces in the North of the Netherlands (Drenthe, Friesland, Groningen). NARSAD Young Investigator Grant from the Brain & Behavior Research Foundation. VENI grant from the Talent Program of the Netherlands Organisation for Scientific Research (NWO-ZonMW 09150161810021). We thank Trynke de Jong for the contribution to Lifelines data collection. We thank Martje Bos and Victoria Trindade Pons for their help in preparing the Lifelines phenotype data. MooDFOOD European Union FP7 funding for MooDFOOD Project Multi-country cOllaborative project on the rOle of Diet, FOod-related behaviour, and Obesity in the prevention of Depression (grant agreement no. 613598). TRAILS Participating centers of the TRacking Adolescents Individual Lives Survey (TRAILS) include the University Medical Center and University of Groningen, the University of Utrecht, the Radboud Medical Center Nijmegen, and the Parnassia Group, all in the Netherlands. TRAILS has been financially supported by various grants from the Netherlands Organization for Scientific Research NWO (Medical Research Council program grant GB-MW 940-38-011; ZonMW Brainpower grant 100-001-004; ZonMw Risk Behavior and Dependence grant 60-60600-97-118; ZonMw Culture and Health grant 261-98-710; Social Sciences Council medium-sized investment grants GB-MaGW 480-01-006 and GB-MaGW 480-07-001; Social Sciences Council project grants GB-MaGW 452-04-314 and GB-MaGW 452-06-004; ZonMw Longitudinal Cohort Research on Early Detection and Treatment in Mental Health Care grant 636340002; NWO large-sized investment grant 175.010.2003.005; NWO Longitudinal Survey and Panel Funding 481-08-013 and 481-11-001; NWO Vici 016.130.002, 453-16-007/2735, and Vi.C.191.021; NWO Gravitation 024.001.003), the Dutch Ministry of Justice (WODC), the European Science Foundation (EuroSTRESS project FP-006), the European Research Council (ERC-2017-STG-757364 and ERC-CoG-2015-681466), Biobanking and Biomolecular Resources Research Infrastructure BBMRI-NL (CP 32), the Gratama foundation, the Jan Dekker foundation, the participating universities, and Accare. Statistical analyses are carried out on the Genetic Cluster Computer (http://www.geneticcluster.org), which is financially supported by the Netherlands Scientific Organization (NWO 480-05-003) along with a supplement from the Dutch Brain Foundation. LASA The Longitudinal Aging Study Amsterdam is largely supported by grants from the Netherlands Ministry of Health, Welfare and Sport, Directorate of Long-Term Care. NQplus NQplus was core funded by ZonMw (ZonMw, Grant 91110030); add-on funding was provided by ZonMW Gezonde Voeding (ZonMw, Grant 115100007), BBMRI (Grant BBMRI-NL RP9 and CP2011-38) and Wageningen University and Research. MOTAR The MOTAR study was funded by NWO VICI grant number 91811602 of B.W.J.H. Penninx. NWO had no role in the design of the study, the collection, analysis and interpretation of the data, or in the preparation, review, or approval of the manuscript. The Hoorn Studies The GWAS in the Hoorn studies was supported by the Amsterdam University Medical Center, a grant from the Foundation for the National Institutes of Health through the Accelerating Medicines Partnership (no. HART17AMP) and the Dutch String of Pearls Initiative. We appreciate the cooperation of the participants and research assistants who have been involved in the Hoorn Study and New Hoorn Study. We would like to thank Tootje Hoovers and Jolanda Bosman as well as all the researchers previously involved for the organization of both studies. Netherlands Twin Register NTR acknowledges funding from the Netherlands Organization for Scientific Research (NWO): Biobanking and Biomolecular Research Infrastructure (BBMRI-NL, 184.033.111) and the BBMRI-NL funded BIOS Consortium (NWO184.021.007); The Netherlands Twin Register is supported by grant NWO 480-15-001/674: Netherlands Twin Registry Repository: researching the interplay between genome and environment, the Avera Institute for Human Genetics and by multiple grants from the Netherlands Organization for Scientific Research (NWO). Genotyping was made possible by grants from NWO/SPI 56-464-14192, Genetic Association Information Network (GAIN) of the Foundation for the National Institutes of Health, Rutgers University Cell and DNA Repository (NIMH U24 MH 068457-06), the Avera Institute, Sioux Falls (USA) and the National Institutes of Health (NIH R01 HD042157-01A1, MH081802, Grand Opportunity grants 1RC2 MH089951 and 1RC2 MH089995) and European Research Council (ERC-230374). DIB acknowledges the Royal Netherlands Academy of Science Professor Award (PAH/6635). Nijmegen Biomedical Study The Nijmegen Biomedical Study is a population-based survey conducted at the Department for Health Evidence and the Department of Laboratory Medicine of the Radboud university medical center. Principal investigators of the Nijmegen Biomedical Study are L.A.L.M. Kiemeney, A.L.M. Verbeek, D.W. Swinkels en B. Franke. Doetinchem Cohort Study The Doetinchem Cohort Study is supported by the Dutch Ministry of Health, Welfare and Sport and the National Institute for Public Health and the Environment. We thank the respondents, epidemiologists and fieldworkers of the Municipal Health Service in Doetinchem for their contribution to the data collection for this study. The authors want to acknowledge the logistic management which was provided by P Vissink, and the data managers J van der Laan, R J de Kleine, I Toxopeus. Further, we thank all (senior) researchers who contributed to the data for collection, in particular in (alphabetical order): J M A de Boer, H B Bueno de Mesquita, P Engelfriet, G C Herber-Gast, G Hulsegge, D Kromhout, L Launer, A C J Nooyens, M C Ocke, S H van Oostrom, K Proper, J C Seidell, H A Smit, W G C Wendel-Vos. NESDA & NESDAsib The infrastructure for the NESDA study (www.nesda.nl) is funded through the Geestkracht program of the Netherlands Organisation for Health Research and Development (ZonMw, grant number 10-0001002) and financial contributions by participating universities and mental health care organizations (VU University Medical Center, GGZ inGeest, Leiden University Medical Center, Leiden University, GGZ Rivierduinen, University Medical Center Groningen, University of Groningen, Lentis, GGZ Friesland, GGZ Drenthe, Rob Giel Onderzoekscentrum). NESDO The infrastructure for NESDO is funded through the Fonds NutsOhra, Stichting tot Steun VCVGZ, NARSAD The Brain and Behaviour Research Fund, and the participating universities and mental health care organizations (VU University Medical Center, Leiden University Medical Center, University Medical Center Groningen, Radboud University Nijmegen Medical Center, and GGZ inGeest, GGNet, GGZ Nijmegen, GGZ Rivierduinen, Lentis, and Parnassia).

## References

1. Marx W, Penninx BWJH, Solmi M, Furukawa TA, Firth J, Carvalho AF, Berk M (2023): Major depressive disorder. Nat Rev Dis Primer 9: 44.

2. Berk M, Köhler-Forsberg O, Turner M, Penninx BWJH, Wrobel A, Firth J, et al. (2023): Comorbidity between major depressive disorder and physical diseases: a comprehensive review of epidemiology, mechanisms and management. World Psychiatry 22: 366–387.

3. American Psychiatric Association (2013): Diagnostic and Statistical Manual of Mental Disorders, 5th ed. Washington, DC: American Psychiatric Publishing.

4. Penninx BWJH, Lamers F, Jansen R, Berk M, Khandaker GM, Picker LD, Milaneschi Y (2025): Immuno-metabolic depression: from concept to implementation. Lancet Reg Health - Eur 48: 101166.

5. Cai N, Choi KW, Fried EI (2020): Reviewing the genetics of heterogeneity in depression: operationalizations, manifestations and etiologies. Hum Mol Genet 29: R10–R18.

6. Milaneschi Y, Lamers F, Penninx BWJH (2021): Dissecting Depression Biological and Clinical Heterogeneity—The Importance of Symptom Assessment Resolution. JAMA Psychiatry 78: 341.

7. Lamers F, Milaneschi Y, Jonge P de, Giltay EJ, Penninx BWJH (2018): Metabolic and inflammatory markers: associations with individual depressive symptoms. Psychol Med 48: 1102–1110.

8. Kim J-M, Kang H-J, Kim J-W, Kim H, Jhon M, Lee J-Y, et al. (2025): Differentiating Subtypes of Major Depressive Disorder Using Serum Biomarkers. J Clin Psychiatry 86. 10.4088/JCP.25m15828

9. Haeringen M van, Milaneschi Y, Lamers F, Penninx BWJH, Jansen R (2023): Dissection of depression heterogeneity using proteomic clusters. Psychol Med 53: 2904–2912.

10. Suseelan S, Pinna G (2023): Heterogeneity in major depressive disorder: The need for biomarker-based personalized treatments. pp 1–67.

11. Nguyen T-D, Harder A, Xiong Y, Kowalec K, Hägg S, Cai N, et al. (2022): Genetic heterogeneity and subtypes of major depression. Mol Psychiatry 27: 1667–1675.

12. Milaneschi Y, Lamers F, Peyrot WJ, Baune BT, Breen G, Dehghan A, et al. (2017): Genetic Association of Major Depression With Atypical Features and Obesity-Related Immunometabolic Dysregulations. JAMA Psychiatry 74: 1214.

13. Musliner KL, Munk-Olsen T, Laursen TM, Eaton WW, Zandi PP, Mortensen PB (2016): Heterogeneity in 10-Year Course Trajectories of Moderate to Severe Major Depressive Disorder. JAMA Psychiatry 73: 346.

14. Eeden WA van, Hemert AM van, Carlier IVE, Penninx BW, Giltay EJ (2019): Severity, course trajectory, and within-person variability of individual symptoms in patients with major depressive disorder. Acta Psychiatr Scand 139: 194–205.

15. Lai W, Liao Y, Zhang H, Zhao H, Li Y, Chen R, et al. (2025): The trajectory of depressive symptoms and the association with quality of life and suicidal ideation in patients with major depressive disorder. BMC Psychiatry 25: 310.

16. Benrimoh D, Kleinerman A, Furukawa TA, III CFR, Lenze EJ, Karp J, et al. (2024): Towards Outcome-Driven Patient Subgroups: A Machine Learning Analysis Across Six Depression Treatment Studies. Am J Geriatr Psychiatry 32: 280–292.

17. Tang H, Xia Y, Gao C, Cai Y, Shao Y, Chen W, et al. (2025): Association of psychosocial factors and biological pathways identified from rare-variant analysis with longitudinal trajectories of treatment response in major depressive disorder. BMC Psychiatry 25: 505.

18. Thomas NS, Gillespie NA, Neale MC, Rosmalen JGM, Loo HM van, Kendler KS (2025): Clinical heterogeneity in major depressive disorder underlies comorbidity with functional disorders. J Psychiatr Res 183: 16–24.

19. Unick GJ, Snowden L, Hastings J (2009): Heterogeneity in Comorbidity Between Major Depressive Disorder and Generalized Anxiety Disorder and Its Clinical Consequences. J Nerv Ment Dis 197: 215–224.

20. Beijers L, Wardenaar KJ, Loo HM van, Schoevers RA (2019): Data-driven biological subtypes of depression: systematic review of biological approaches to depression subtyping. Mol Psychiatry 24: 888–900.

21. Beckett CW, Lawson RP (2026): From symptoms to systems: A comprehensive review of the clinical utility of classic and novel approaches to major depression subtyping. J Affect Disord 395: 120731.

22. Flint J (2023): The genetic basis of major depressive disorder. Mol Psychiatry 28: 2254–2265.

23. Huider F, Milaneschi Y, van der Zee MD, de Geus EJC, Helmer Q, Penninx BWJH, Boomsma DI (2021): Major Depressive Disorder and Lifestyle: Correlated Genetic Effects in Extended Twin Pedigrees. Genes 12: 1509.

24. Meng X, Navoly G, Giannakopoulou O, Levey DF, Koller D, Pathak GA, et al. (2024): Multi-ancestry genome-wide association study of major depression aids locus discovery, fine mapping, gene prioritization and causal inference. Nat Genet 56: 222–233.

25. Adams MJ, Streit F, Meng X, Awasthi S, Adey BN, Choi KW, et al. (2025): Trans-ancestry genome-wide study of depression identifies 697 associations implicating cell types and pharmacotherapies. Cell 188: 640–652.e9.

26. Mitchell BL, Monistrol-Mula A, Thomas JT, Byrne EM (2025): The Role of Genetic Data in Dissecting Depression Heterogeneity. Biol Psychiatry. 10.1016/j.biopsych.2025.10.029

27. Thorp JG, Marees AT, Ong J-S, An J, MacGregor S, Derks EM (2020): Genetic heterogeneity in self-reported depressive symptoms identified through genetic analyses of the PHQ-9. Psychol Med 50: 2385–2396.

28. Shorter JR, Pasman JA, Kurvits S, Jangmo A, Naamanka J, Harder A, et al. (2025): Genome-wide association analyses identify distinct genetic architectures for early-onset and late-onset depression. Nat Genet 57: 2972–2979.

29. Balding DJ, Speed D (2025): Recent Statistical Innovations in Human Genetics. Ann Hum Genet 89: 241–254.

30. Grotzinger AD, Rhemtulla M, Vlaming R de, Ritchie SJ, Mallard TT, Hill WD, et al. (2019): Genomic structural equation modelling provides insights into the multivariate genetic architecture of complex traits. Nat Hum Behav 3: 513–525.

31. Adams MJ, Thorp JG, Jermy BS, Kwong ASF, Kõiv K, Grotzinger AD, et al. (2024): Genome-wide meta-analysis of ascertainment and symptom structures of major depression in case-enriched and community cohorts. Psychol Med 54: 3459–3468.

32. Gilchrist L, Spargo TP, Green RE, Coleman JRI, Howard DM, Thorp JG, et al. (2025): Depression symptom-specific genetic associations in clinically diagnosed and proxy case Alzheimer’s disease. Nat Ment Health 3: 212–228.

33. Huider F, Milaneschi Y, Hottenga J-J, Bot M, Rietman ML, Kok AAL, et al. (2024): Genomics Research of Lifetime Depression in the Netherlands: The BIObanks Netherlands Internet Collaboration (BIONIC) Project. Twin Res Hum Genet 27: 1–11.

34. R Core Team (2024): R: A language and environment for statistical computing (Version 4.4.2). R Foundation for Statistical Computing. Retrieved from https://www.r-project.org/

35. Python Software Foundation (2024): Python (Version 3.11). Retrieved from https://www.python.org/

36. Huider F, Milaneschi Y, Pool R, Maciel B de APC, Gordon SD, Rietman ML, et al. (2025): Genetics of Major Depressive Disorder in a Homogeneous Population with Uniform Phenotyping. 10.1101/2025.05.14.25325937

37. Jiang L, Zheng Z, Qi T, Kemper KE, Wray NR, Visscher PM, Yang J (2019): A resource-efficient tool for mixed model association analysis of large-scale data. Nat Genet 51: 1749–1755.

38. Yang J, Lee SH, Goddard ME, Visscher PM (2011): GCTA: A Tool for Genome-wide Complex Trait Analysis. Am J Hum Genet 88: 76–82.

39. Harder A (2025): ldsR. Retrieved from https://github.com/Ararder/ldsR

40. Bulik-Sullivan BK, Loh P-R, Finucane HK, Ripke S, Yang J, Patterson N, et al. (2015): LD Score regression distinguishes confounding from polygenicity in genome-wide association studies. Nat Genet 47: 291–295.

41. Peyrot WJ, Price AL (2021): Identifying loci with different allele frequencies among cases of eight psychiatric disorders using CC-GWAS. Nat Genet 53: 445–454.

42. Auton A, Abecasis GR, Altshuler DM, Durbin RM, Abecasis GR, Bentley DR, et al. (2015): A global reference for human genetic variation. Nature 526: 68–74.

43. Ning Z (2025): HDL. Retrieved from https://github.com/zhenin/HDL

44. Ning Z, Pawitan Y, Shen X (2020): High-definition likelihood inference of genetic correlations across human complex traits. Nat Genet 52: 859–864.

45. Have M ten, Tuithof M, Dorsselaer S van, Schouten F, Luik AI, Graaf R de (2023): Prevalence and trends of common mental disorders from 2007-2009 to 2019-2022: results from the Netherlands Mental Health Survey and Incidence Studies (NEMESIS), including comparison of prevalence rates before vs. during the COVID-19 pandemic. World Psychiatry 22: 275–285.

46. Bycroft C, Freeman C, Petkova D, Band G, Elliott LT, Sharp K, et al. (2018): The UK Biobank resource with deep phenotyping and genomic data. Nature 562: 203–209.

47. Davies MR, Kalsi G, Armour C, Jones IR, McIntosh AM, Smith DJ, et al. (2019): The Genetic Links to Anxiety and Depression (GLAD) Study: Online recruitment into the largest recontactable study of depression and anxiety. Behav Res Ther 123: 103503.

48. Creese B, Arathimos R, Brooker H, Aarsland D, Corbett A, Lewis C, et al. (2021): Genetic risk for Alzheimer’s disease, cognition, and mild behavioral impairment in healthy older adults. Alzheimers Dement Diagn Assess Dis Monit 13. 10.1002/dad2.12164

49. Zaykin DV, Kozbur DO (2010): P-value based analysis for shared controls design in genome-wide association studies. Genet Epidemiol 34: 725–738.

50. Fried EI, Nesse RM (2015): Depression is not a consistent syndrome: An investigation of unique symptom patterns in the STAR*D study. J Affect Disord 172: 96–102.

51. Pistis G, Strippoli M-PF, Dalfsen JH van, Vaucher J, Kutalik Z, Vollenweider P, et al. (2025): The Role of Energy Homeostasis in Depression Pathophysiology and Its Heterogeneity. JAMA Psychiatry 82: 992.

52. Cosgrove KT, Burrows K, Avery JA, Kerr KL, DeVille DC, Aupperle RL, et al. (2020): Appetite change profiles in depression exhibit differential relationships between systemic inflammation and activity in reward and interoceptive neurocircuitry. Brain Behav Immun 83: 163–171.

53. Kroemer NB, Opel N, Teckentrup V, Li M, Grotegerd D, Meinert S, et al. (2022): Functional Connectivity of the Nucleus Accumbens and Changes in Appetite in Patients With Depression. JAMA Psychiatry 79: 993.

54. Simmons WK, Burrows K, Avery JA, Kerr KL, Bodurka J, Savage CR, Drevets WC (2016): Depression-Related Increases and Decreases in Appetite: Dissociable Patterns of Aberrant Activity in Reward and Interoceptive Neurocircuitry. Am J Psychiatry 173: 418–428.

55. Simmons WK, Burrows K, Avery JA, Kerr KL, Taylor A, Bodurka J, et al. (2020): Appetite changes reveal depression subgroups with distinct endocrine, metabolic, and immune states. Mol Psychiatry 25: 1457–1468.

56. Kendler KS, Aggen SH, Neale MC (2013): Evidence for multiple genetic factors underlying DSM-IV criteria for major depression. JAMA Psychiatry 70: 599–607.

57. Van Loo HM, Aggen SH, Kendler KS (2022): The structure of the symptoms of major depression: Factor analysis of a lifetime worst episode of depressive symptoms in a large general population sample. J Affect Disord 307: 115–124.

58. Fanelli G, Franke B, Witte WD, Ruisch IH, Haavik J, Gils V van, et al. (2022): Insulinopathies of the brain? Genetic overlap between somatic insulin-related and neuropsychiatric disorders. Transl Psychiatry 12: 59.

59. Fanelli G, Franke B, Fabbri C, Werme J, Erdogan I, Witte WD, et al. (2025): Local patterns of genetic sharing between neuropsychiatric and insulin resistance-related conditions. Transl Psychiatry 15: 145.

60. Badini I, Coleman JRI, Hagenaars SP, Hotopf M, Breen G, Lewis CM, Fabbri C (2022): Depression with atypical neurovegetative symptoms shares genetic predisposition with immuno-metabolic traits and alcohol consumption. Psychol Med 52: 726–736.

61. Brydges CR, Bhattacharyya S, Dehkordi SM, Milaneschi Y, Penninx B, Jansen R, et al. (2022): Metabolomic and inflammatory signatures of symptom dimensions in major depression. Brain Behav Immun 102: 42–52.

62. Bergstedt J, Pasman JA, Ma Z, Harder A, Yao S, Parker N, et al. (2024): Distinct biological signature and modifiable risk factors underlie the comorbidity between major depressive disorder and cardiovascular disease. Nat Cardiovasc Res 3: 754–769.

63. Alshehri T, Mook-Kanamori DO, Willems van Dijk K, Dinga R, Penninx BWJH, Rosendaal FR, et al. (2023): Metabolomics dissection of depression heterogeneity and related cardiometabolic risk. Psychol Med 53: 248–257.

64. Milaneschi Y, Lamers F, Berk M, Penninx BWJH (2020): Depression Heterogeneity and Its Biological Underpinnings: Toward Immunometabolic Depression. Biol Psychiatry 88: 369–380.

65. Lamers F, Milaneschi Y, Vinkers CH, Schoevers RA, Giltay EJ, Penninx BWJH (2020): Depression profilers and immuno-metabolic dysregulation: Longitudinal results from the NESDA study. Brain Behav Immun 88: 174–183.

66. Lamers F, Bot M, Jansen R, Chan MK, Cooper JD, Bahn S, Penninx BWJH (2016): Serum proteomic profiles of depressive subtypes. Transl Psychiatry 6: e851.

67. Hagenberg J, Brückl TM, Erhart M, Kopf-Beck J, Ködel M, Rehawi G, et al. (2024): Dissecting depression symptoms: Multi-omics clustering uncovers immune-related subgroups and cell-type specific dysregulation. Brain Behav Immun 123: 353–369.

68. Sforzini L, Marizzoni M, Bottanelli C, Kunšteková V, Zonca V, Saleri S, et al. (2025): Transcriptomic profiles in major depressive disorder: the role of immunometabolic and cell-cycle-related pathways in depression with different levels of inflammation. Mol Psychiatry 30: 1308–1318.

69. Vreijling SR, Haeringen M van, Milaneschi Y, Huider F, Bot M, Amin N, et al. (2023): Sociodemographic, lifestyle and clinical characteristics of energy-related depression symptoms: A pooled analysis of 13,965 depressed cases in 8 Dutch cohorts. J Affect Disord 323: 1–9.

70. Lamers F, Jonge P de, Nolen WA, Smit JH, Zitman FG, Beekman ATF, Penninx BWJH (2010): Identifying depressive subtypes in a large cohort study: results from the Netherlands Study of Depression and Anxiety (NESDA). J Clin Psychiatry 71: 1582–9.

71. Rydin AO, Milaneschi Y, Lamers F, Quax R, Bunt N van de, Koloi A, et al. (2025): Trajectories of depressive symptoms, metabolic syndrome, inflammation, and cardiometabolic diseases: A longitudinal Bayesian network approach. Brain Behav Immun 130: 106120.

72. Toenders YJ, Schmaal L, Harrison BJ, Dinga R, Berk M, Davey CG (2020): Neurovegetative symptom subtypes in young people with major depressive disorder and their structural brain correlates. Transl Psychiatry 10: 108.

73. Toenders YJ, Schmaal L, Nawijn L, Han LKM, Binnewies J, Wee NJA van der, et al. (2022): The association between clinical and biological characteristics of depression and structural brain alterations. J Affect Disord 312: 268–274.

74. Tang L, Tang R, Zheng J, Zhao P, Zhu R, Tang Y, et al. (2025): Dissecting biological heterogeneity in major depressive disorder based on neuroimaging subtypes with multi-omics data. Transl Psychiatry 15: 72.

75. Wang Q, Dwivedi Y (2025): Recent developments in omics studies and artificial intelligence in depression and suicide. Transl Psychiatry 15: 275.

76. Sathyanarayanan A, Mueller TT, Moni MA, Schueler K, Baune BT, Lio P, et al. (2023): Multi-omics data integration methods and their applications in psychiatric disorders. Eur Neuropsychopharmacol 69: 26–46.

77. Cai N, Revez JA, Adams MJ, Andlauer TFM, Breen G, Byrne EM, et al. (2020): Minimal phenotyping yields genome-wide association signals of low specificity for major depression. Nat Genet 52: 437–447.

78. Sullivan MLC, Schwaba T, Harden KP, Grotzinger AD, Nivard MG, Tucker-Drob EM (2024): Beyond the factor indeterminacy problem using genome-wide association data. Nat Hum Behav 8: 205–218.

