## Supplemental Materials for "Symptom-specific genetics reveal heterogeneity within major depressive disorder"

### S0. BIONIC

The BIONIC project is a nation-wide initiative harmonizing the assessment of lifetime major depressive disorder (MDD) across 16 Dutch cohorts (*N*=132,850), with genome-wide SNP data currently available for 66,684 participants. Case–control definitions were based on DSM-5 criteria, assessed primarily through the Lifetime Depression Assessment Survey (LIDAS)(1), an online self-report instrument adapted from the CIDI-SF (2) and comprising information on demographics/lifestyle, depressive symptoms, and diagnostic history. Supplementary diagnostic data from structured interviews (CIDI and MINI (3)) were incorporated when compatible with DSM-V definitions.

MDD was defined as the presence of at least five out of nine DSM-V depression symptoms (including at least one cardinal symptom), accompanied by significant impairment lasting a minimum of two weeks. Controls were defined as individuals with fewer than five symptoms, no cardinal symptoms, or symptom duration/impairment too short to meet diagnostic thresholds (<2 weeks). Additional control subjects who did not complete LIDAS were identified using conservative thresholds on validated self-report measures (e.g., Beck Depression Inventory (4)). Controls were screened for a diagnostic or treatment history of depression, bipolar disorder, schizophrenia, obsessive-compulsive disorder, post-traumatic stress disorder, phobia, attention deficit hyperactivity disorder/attention deficit disorder, as well as any personality, eating, or substance use disorder. Any such condition resulted in exclusion from the control group.

### S1. Individual Symptom GWAS

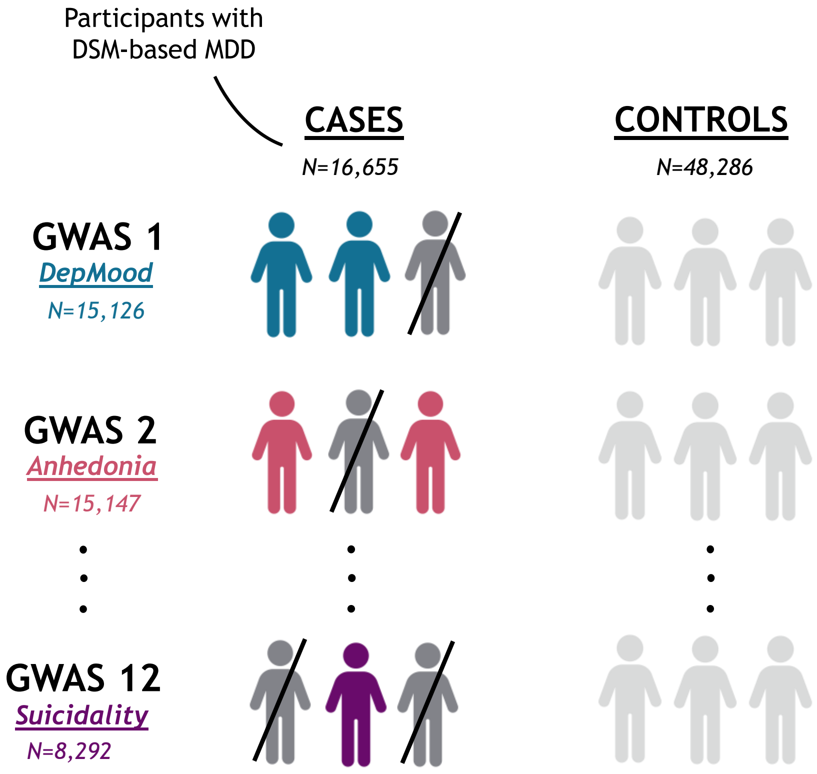


**SFigure 1**. Schematic representation of the twelve symptom-level Genome-Wide Association Studies (GWAS) conducted in BIONIC. Participants with and without DSM-based MDD were previously defined by Huider et al (5).

#### S1.1 GWAS Diagnostics

SNPs with low imputation quality (INFO score < 0.6) and minor allele frequency (MAF) < 0.01 were filtered out and excluded from further analyses. Genomic inflation factors ($\lambda$ and its standardized measure $\lambda_{1000}$) as well as the intercept from Linkage Disequilibrium Score Regression (LDSC) (6) were calculated to assess potential inflation of test statistics due to population stratification or other confounding factors (S1.1) (6,7). Lamda ($\lambda$) represents the median observed chi-square statistic divided by the expected median under the null hypothesis, providing a measure of overall test statistic inflation (7). To allow comparisons across studies of different sizes, $\lambda_{1000}$ standardizes the $\lambda$ measure to a sample size of 1,000 individuals (8). Values of $\lambda$ or $\lambda_{1000}$ above 1 may indicate inflation due to population stratification, cryptic relatedness, or uncontrolled confounding, whereas values close to 1 suggest well-calibrated test statistics (9).

#### S1.2 Significant SNPs

The filtered summary statistics were also annotated via *FUMA$^{GWAS}$* (version 1.6.3) (40). In total, we identified three significant independent risk loci, namely *rs3818852* for Depressed Mood and *rs147067514* for Psychomotor Agitation. Those SNPs were annotated via *FUMA* to 7 and 4 genes, respectively. More information on the annotated genes can be found in (ST2a).

Note that the SNP significance results should be interpreted with caution, as using the same control group across multiple symptom-level GWAS may introduce biases. These may distort the p-values and lead to over- or under-estimation of SNP associations (10). Further validation in independent datasets or with separate control groups for each symptom may help to confirm the robustness of these findings.

### SFigure 2. Manhattan and QQ plots for the symptom GWAS with significant hits

- For Depressed Mood

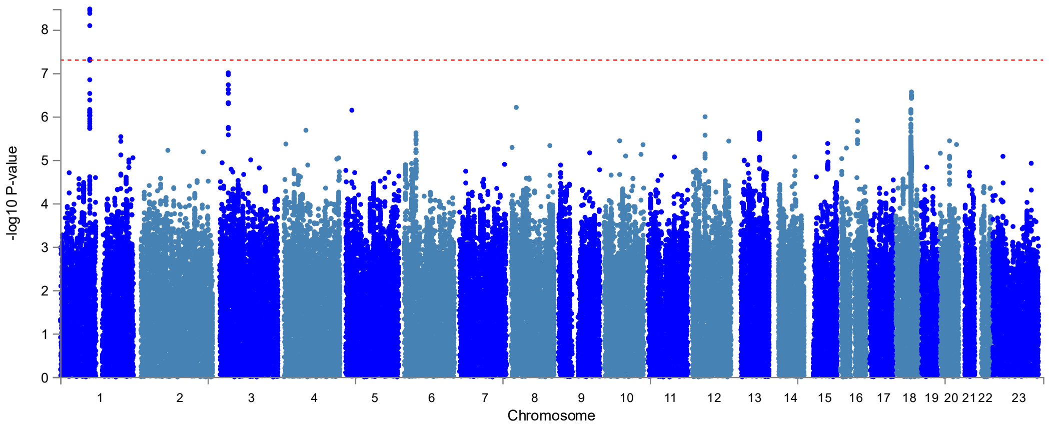


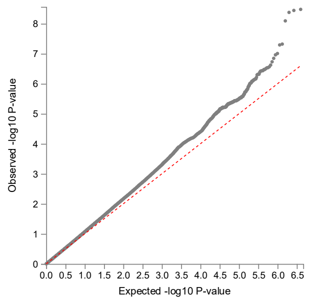


- For Psychomotor Agitation

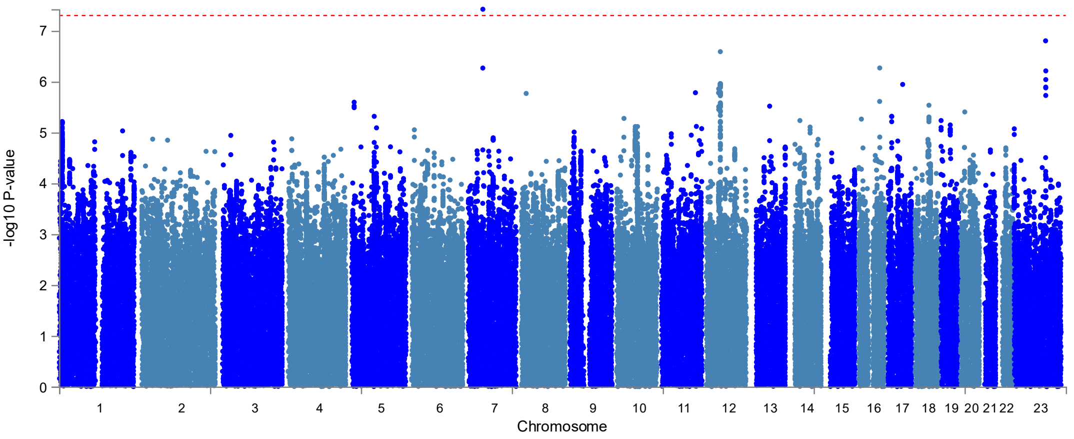


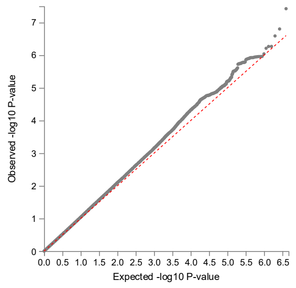

##### S1.3 Functional Annotation

The significantly associated genes were functionally annotated using the *GENE2FUNC* pipeline of *FUMA*. This includes enrichment gene set analyses as well as expression quantitative trait loci (eQTL) analyses using *GTEx v8 54 tissue types* and *GTEx v8 30 general tissue types* (11).

The enrichment analyses revealed distinct gene set associations for both loci (ST2b). SNP *rs3818852* was linked to two GWAS catalog gene sets related to incident chronic kidney disease and white matter integrity in the context of bipolar disorder. SNP *rs147067514* was associated with a GWAS catalog gene set linked to gut microbiota alpha diversity, as measured by the Shannon index.

##### S1.4 Heritabilities

To calculate heritability *Z*-scores, we divided the $h^2_{SNP}$ estimates by their standard error for each individual trait. Corresponding p-values were derived from the *Z*-scores assuming a two-sided standard normal distribution.

#### S2. Identification of underlying factors

##### S2.1 Chromosome-Split Analysis

To avoid overfitting during the common factor analysis, we attempted to split the summary statistics by odd and even chromosomes, as is common practice in genomic structural equation modelling (GenomicSEM) studies of highly polygenic traits (12–14). This approach involves performing exploratory factor analysis (EFA) on odd chromosomes and validating the results using confirmatory factor analysis (CFA) on even chromosomes, or vice versa. However, in our symptom GWAS data, heritability was unevenly distributed between odd and even chromosomes (ST4a, ST4b), likely indicating a sparse genetic architecture with relatively stronger effects concentrated in specific loci-chromosomes. As this resulted in unstable and unreliable model solutions, we did not pursue this approach further and proceeded with the full set of summary statistics instead.

#### S2.2 HDL Estimation for GenomicSEM

Compared to the standard Linkage Disequilibrium Score Regression (LDSC), HDL yields more precise estimates by reducing the variance of each genetic correlation by ~60% (15). The reference panel consisted of 1,029,876 quality-controlled, imputed HapMap3 SNPs from the UK Biobank (16,17).

In Genomic SEM, as a first step, we applied the *munge* function to the raw GWAS summary statistics, using the default parameters (MAF ≥ 0.01; imputation INFO score > 0.9) and HapMap3 as the reference panel. To obtain a genetic correlation matrix compatible with the input formatting requirements of GenomicSEM, we re-ran HDL within the GenomicSEM framework. We used HapMap3 as the reference panel and selected the “piecewise” method.

#### S2.3 Prioritizing Sub-Symptoms for Model Specificity

The pairwise genetic correlations between symptoms did not include the broad phenotypes Appetite/Weight Changes, Sleep Problems, or Psychomotor Changes, rather than their specific sub-symptoms (e.g. Appetite/Weight Gain vs Loss). Prioritizing sub-symptoms (e.g., appetite or weight gain vs. loss) over broad phenotypes enabled a more granular representation of depressive symptomatology and avoided complicating the model with overlapping constructs.

#### S2.4 Prevalence Rescaling

To run HDL within Genomic SEM, we rescaled the symptom prevalences observed among MDD cases in the BIONIC dataset by the estimated population prevalence of lifetime MDD in the Netherlands (~25%) (18). This adjustment was necessary because we aim to approximate population-level prevalences of each symptom conditional on an MDD diagnosis —information that, to our knowledge, is not directly available from existing observational studies.

#### S2.5 EFA Rotation Strategy

We first employed the rotation "*varimax*" (orthogonal method) (19) but, due to poor model fit and low discriminatory power, we switched to rotation "*promax*", as implemented in numerous Genomic SEM studies (20–23). This technique assumes oblique axes, allowing correlated factors and markedly improving model fit. We therefore proceeded with this rotation in all reported models.

#### S2.6 CFA Model Specifications

During confirmatory factor analysis (CFA), a factor loading threshold of 0.3 was applied, with no restrictions on the number of symptoms allowed to load onto each factor. Cross-loadings were also permitted (i.e., symptoms loading onto more than one factor), as excluding them did not enhance model fit and occasionally even compromised it.

#### S2.7 Model Evaluation and Factor Retention

For the assessment of model performance, we consulted several fit indices for each EFA-derived model (STable1): Comparative Fit Index (*CFI*), Standardized Root Mean Square Residual (*SRMR*), Akaike Information Criterion (*AIC*), and the chi-square statistic (*Chisq*).

**STable 1.** Performance metrics per model. *CFI* ≥ 0.95 (excellent), ≥ 0.90 (acceptable); *SRMR* ≤ 0.05 (excellent), ≤ 0.08 (acceptable).

| | *CFI* | *SRMR* | *AIC* | *Chisq* |
|---|---|---|---|---|
| **Model 1** | 0.815 | 0.048 | 9541037 | 9540989 |
| **Model 2** | 0.894 | 0.039 | 5455366 | 5455312 |
| **Model 3** | 0.944 | 0.032 | 2881593 | 2881533 |
| **Model 4** | 0.886 | 0.043 | 5884526 | 5884464 |
| **Model 5** | 0.908 | 0.036 | 728021 | 727961 |

To assist in determining the optimal number of factors, we conducted parallel analysis by simulating random genetic correlation matrices and comparing their eigenvalues to those of the observed data (SFigure1). By taking into consideration the suggested number of factors along with the model performance metrics, we aimed to balance the trade-off between model fit and parsimony. Factors were retained if their eigenvalues from the observed data were greater than the corresponding eigenvalues from the simulated data (24). Additionally, we considered the Kaiser criterion, which suggests retaining factors with eigenvalues greater than 1 (25).

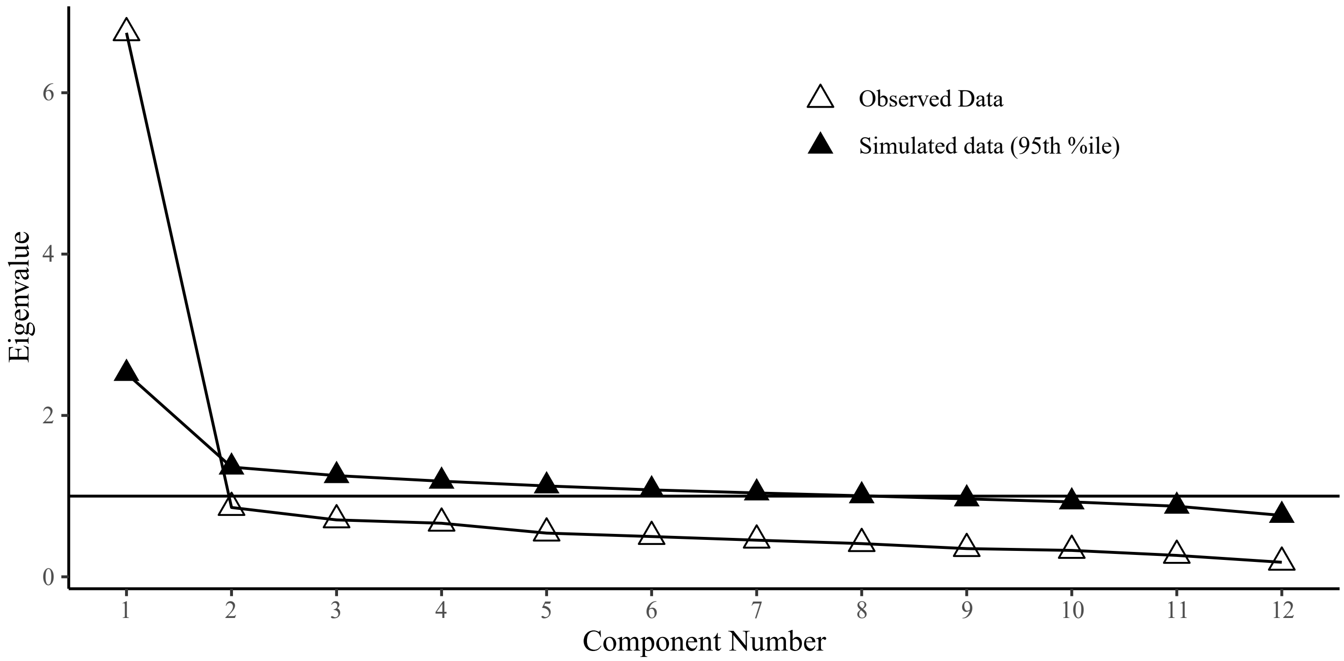


**SFigure 3.** Scree plot from parallel analysis involving observed and simulated eigenvalues. *Component Number = Number of Factors*

In our case, the optimal number of factors was identified prior to the two-factor solution, suggesting a strong general genetic factor underlying depressive symptoms. Nevertheless, to allow for some degree of discrimination and the possibility of identifying more specific genetic entities within depression, we selected the two-factor solution for further analyses. The model's full results, along with the other EFA-derived models can be found in Supplementary Data ST5a-e, while STable2 summarizes each model's factor composition as determined by the EFA.

**STable 2.** Factor composition of depressive symptoms, as indicated by exploratory factor analyses for 1-5 factors. (-) indicates negative loadings.

| | F1 | F2 | F3 | F4 | F5 |
|---|---|---|---|---|---|
| **Model 1** | All depressive symptoms | - | - | - | - |
| **Model 2** | Fatigue<br>AppWLoss<br>Insomnia<br>Hypersomnia<br>Slowing<br>Agitation<br>Guilt/Worth<br>Suicidality | DepMood<br>Anhedonia<br>Fatigue<br>AppWGain<br>Agitation<br>ConcProb | - | - | - |
| **Model 3** | Fatigue<br>AppWGain<br>Insomnia<br>Hypersomnia<br>Slowing<br>Agitation<br>Guilt/Worth<br>Suicidality | DepMood<br>Anhedonia<br>Fatigue<br>ConcProb | AppWLoss<br>(-)AppWGain<br>Hypersomnia | - | - |
| **Model 4** | DepMood<br>Anhedonia<br>Fatigue<br>AppWGain<br>Hypersomnia<br>Slowing | Insomnia | Guilt/Worth | AppWLoss<br>(-)AppWGain | - |

| | | | | | |
|---|---|---|---|---|---|
| | Agitation ConcProb Suicidality | | | | |
| **Model 5** | DepMood Anhedonia Fatigue AppWGain, ConcProb | AppWGain Insomnia Agitation Suicidality | Guilt/Worth | Hypersomnia | AppWLoss |

#### S2.8 Fitting bifactor and hierarchical models

We also attempted to fit bifactor and hierarchical models based on the symptom-factor structure identified in the EFA, aiming to better capture the underlying genetic architecture. However, these more constrained models could not be successfully fitted, possibly due to low genetic variation (within-cases symptoms' genetic correlations).

### S3. Factor Assessment

#### S3.1 External Phenotypes for Comparability Assessment

To assess construct comparability and replication potential, we compared the symptom definitions in BIONIC with those of two external datasets that, to our knowledge, best approximate our phenotypic traits. In BIONIC, cases were defined as individuals with a clinical diagnosis of MDD who also endorsed a specific symptom, requiring both diagnostic confirmation and symptom-level endorsement. In the meta-analysis (UKB_meta) by Gilchrist et al. (2025) (26), symptom endorsement was not examined within the context of MDD. Additionally, several symptoms were assessed using non-directional, composite categories (e.g. Appetite/Weight Changes), capturing a broader, less specific phenotype. On the other hand, the PGC (2024) (27) employed symptom definitions comparable in granularity to BIONIC (e.g. distinguishing between Insomnia and Hypersomnia), but defined cases based solely on symptom endorsement, i.e. without requiring an MDD diagnosis. Similarly, controls were defined as participants not endorsing the symptom in question, but were not screened for MDD (PGC_ca: MDD-enriched cohorts endorsing the symptom vs MDD-enriched cohorts not endorsing the symptom-resembling a “case-case” GWAS, PGC_com: community-based cohorts endorsing the symptom vs community-based cohorts not endorsing the symptom). This latter approach likely reduces the specificity of the genetic signal for depression-related symptoms and introduces noise, as such symptoms may arise from other psychiatric or even somatic conditions.

This comparison highlights key conceptual overlaps and definitional differences that are critical for interpreting replication results. Variation in case criteria and symptom granularity plays a crucial role in shaping between-symptom genetic correlations and, consequently, the success of model replication.

#### S3.2 Genetic Correlations for data comparability assessment

We examined genetic correlations between individual depressive symptoms in BIONIC and -when available- their counterparts in UKB_meta and PGC (STable3) in order to assess data comparability across studies. It is worth noting that, of the 12 PGC symptoms, only 7 and 10 were taken forward for PGC_ca and PGC_com, respectively (STable 3). For PGC_ca, the symptoms DepMood, Anhedonia, Fatigue and ConcProb were excluded because of negative heritabilities (likely indicating inadequate power due to low variation), while we also excluded Psychomotor Slowing, as its SNP-based heritability was almost 0. For PGC_com, the symptoms of Psychomotor Slowing and Agitation were excluded, as they did not meet the sample size inclusion criteria ($N_{eff}$ > 5000).

**STable 3.** Genetic correlations between BIONIC depressive symptoms and their counterparts in a) PGC_ca ($N_{eff}$ = 2,471 - 11,130, ST6a), b) PGC_com ($N_{eff}$ = 218 - 102,071, ST6b) and c) UKB_meta ($N_{eff}$ = 224,535 – 308,421, ST7). Estimates involving symptoms with insufficient $N_{eff}$ (< 5,000) or near-zero/negative SNP-based heritability are not interpretable and therefore are not reported.

| | PGC_ca | PGC_com | UKB_meta |
|---|---|---|---|
| **1.** Depressed Mood | - | 0.5578 | 0.7078 |
| **2.** Anhedonia | - | 0.7508 | 0.6577 |
| **3a.** Appetite Decrease or Weight Loss | 0.2175 | 0.4343 | |
| **3b.** Appetite Increase or Weight Gain | 0.5321 | 0.5468 | |
| **4a.** Insomnia | 0.223 | 0.6074 | |
| **4b.** Hypersomnia | 0.5627 | 0.7732 | |
| **5a.** Psychomotor Slowing | - | - | |
| **5b.** Psychomotor Agitation | 0,4584 | - | |
| **6.** Energy Loss / Fatigue | - | 0.5111 | 0.6186 |
| **7.** Feelings of Worthlessness or Guilt | 0.9452 | 0.5404 | 0.6333 |
| **8.** Concentration Problems | - | 0.7083 | 0.6661 |
| **9.** Thoughts about Death or Suicide | 0.4871 | 0.6396 | 0.7745 |

#### S3.3 The Importance of Phenotypic Granularity

Correlating the BIONIC summary statistics with those of UK_meta also offered interesting insights into the importance of phenotype granularity for interpreting symptom-level associations. Focusing on Appetite/Weight Changes, the phenotype by UKB_meta showed the strongest genetic correlation with Appetite/Weight Gain in BIONIC, followed by the broader Appetite/Weight Change, and finally Appetite/Weight Loss (SFigure 4, ST7). This pattern suggests that the genetic signal captured by the broad UKB_meta measure was largely driven by individuals with appetite or weight gain, likely reflecting an overrepresentation of such individuals in their sample. Importantly, this finding underscores the value of phenotypic specificity: collapsing directionally distinct symptoms into a single broad category can cancel out meaningful genetic differences, reducing interpretability and potentially biasing associations.

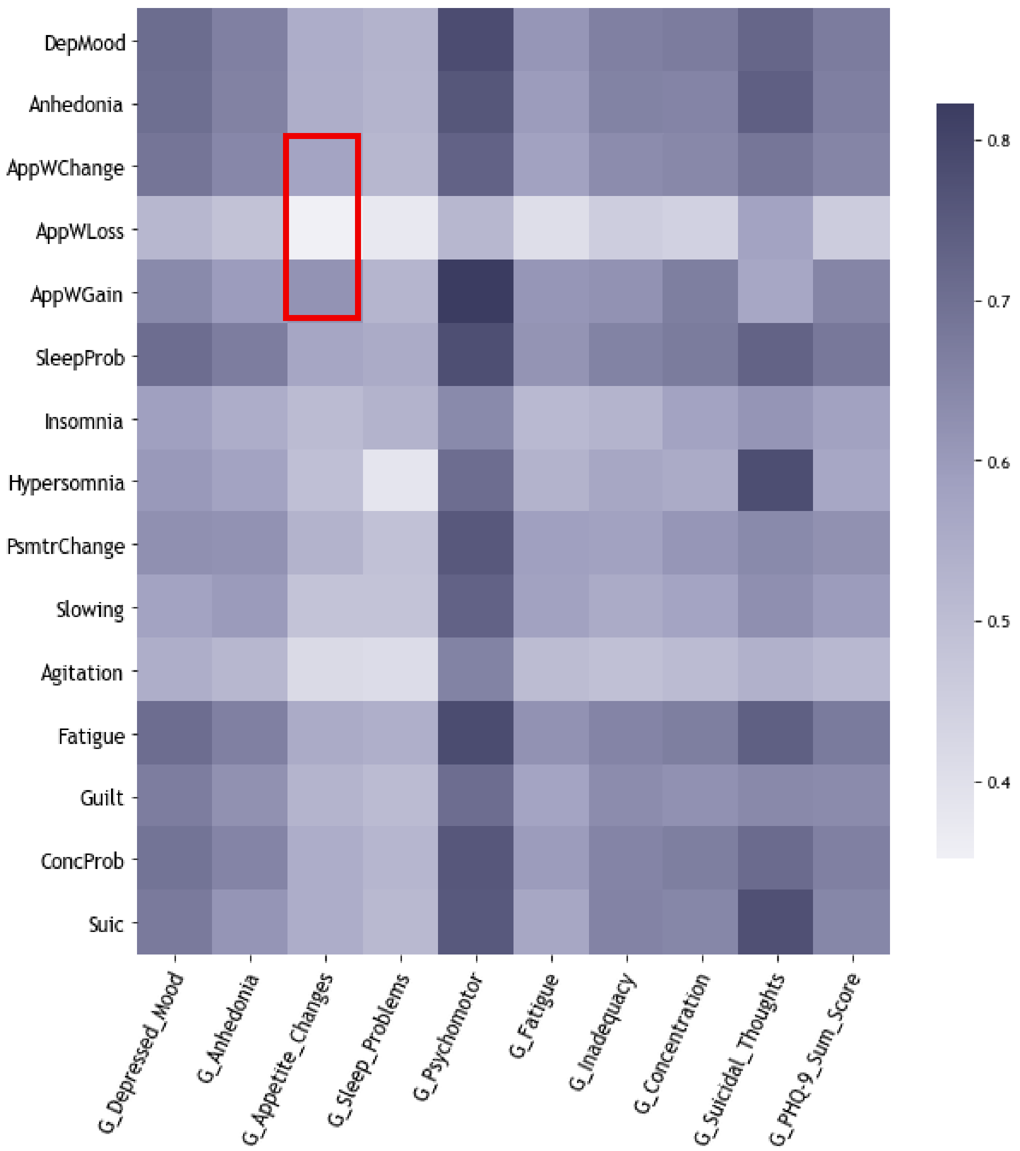


**SFigure 4.** Genetic correlations between the symptoms of BIONIC and those used by Gilchrist et al (G_*). Correlation estimates ranged from 0.35 to 0.82 and were all significant ($p_{fdr}$<0.05).

#### S3.4 Attempting Model Replication

Despite differences in case definition, the finer-grained symptom definitions used by PGC (2024) allowed for a closer approximation of our model in both case-enriched and community-based datasets. However, model fit was suboptimal in both datasets (case-enriched: *CFI*=0.456, *SRMR*=0.22, *AIC*=68793403, *Chisq*=68793349; community-based: *CFI*=0.777, *SRMR*=0.136, *AIC*=3858321, *Chisq*=3858267).

When conducting EFA in the case-enriched dataset, some factor loadings exceeded 1 (STable4), indicating model estimation issues that limit the interpretability of the results and caution against direct structural comparison with other models. In the community-based cohort, the EFA yielded a factor solution largely different from BIONIC, with one emotional-cognitive depression factor and a second factor largely defined by somatic symptoms (STable 4). These findings suggest that while symptom granularity enables more faithful model replication, differences in case ascertainment and population characteristics may still meaningfully affect factor structure.

**STable 4.** Two-factor Exploratory Factor Analysis in the case-enriched and community-based cohorts from PGC (2024).

| | **Case-enriched** | | **Community-based** | |
|---|---|---|---|---|
| | **F1** | **F2** | **F1** | **F2** |
| **S1.** DepMood | 0.129 | - | 0.630 | - |
| **S2.** Anhedonia | 0.217 | - | 0.873 | - |
| **S3a.** AppWLoss | - | 0.415 | 0.222 | 0.638 |
| **S3b.** AppWGain | 0.447 | - | 0.306 | 0.314 |
| **S4a.** Insomnia | -0.275 | 1.026* | 0.559 | 0.643 |
| **S4b.** Hypersomnia | 1.029* | -0.428 | 0.746 | 0.509 |
| **S5a.** Slowing | 0.777 | 0.315 | -0.119 | 0.373 |
| **S5b.** Agitation | -0.100 | 0.725 | 0.161 | -0.314 |
| **S6.** Fatigue | 0.998 | -0.121 | 0.920 | - |
| **S7.** Guilt/Worth | 0.474 | 0.312 | 0.865 | -0.171 |
| **S8.** ConcProb | 0.593 | 0.608 | 0.943 | 0.172 |
| **S9.** Suicidality | 0.543 | - | 0.484 | 0.148 |
| **F1~F2** | 0.361 | | 0.237 | |

### S4. Factor Characterization

#### S4.1 External Phenotypes for Characterization

To conceptualize our model structure, we ran multivariate genetic regression (mvReg) of numerous external traits on our two factors within the GenomicSEM framework. STables 5 and 6 give more information on the GWAS used to examine such phenotypes.

**STable 5.** GWAS of the neuropsychiatric external traits examined in the multivariate regression within GenomicSEM.

| Trait | Abbreviation | Study | PubMedID | *Neff* | Population prevalence used | Accession nr / Website |
|---|---|---|---|---|---|---|
| Attention-Deficit/Hyper-activity Disorder | ADHD | Demontis et al. (2023) (28) | 36702997 | 128,214 | 0.025 | https://doi.org/10.6084/m9.figshare.22564390 |
| Autism Spectrum Disorder | ASD | Grove et al. (2019) (29) | 30804558 | 44,367 | 0.012 | https://doi.org/10.6084/m9.figshare.14671989 |
| Bipolar Disorder | BD | O'Connell et al. (2025) (30) | 39843750 | 191,104 | 0.012 | https://doi.org/10.6084/m9.figshare.27216117 |
| Major Depressive Disorder | MDD | Adams et al. (2025) (31) | 39814019 | 2,008,918 | 0.15 | https://doi.org/10.6084/m9.figshare.27061255 |
| Schizophrenia | SCZ | Trubetskoy et al. (2022) (32) | 35396580 | 146,378 | 0.01 | https://doi.org/10.6084/m9.figshare.19426775 |

**STable 6.** GWAS of the cardiometabolic external traits examined in mvReg.

| Trait | Abbreviation | Binary/Cont. | Study | PubMedID | *N* or *Neff* | Population prevalence used | Accession nr / Website |
|---|---|---|---|---|---|---|---|
| Body Mass Index | BMI | Continuous | Yengo et al. (2018) (33) | 30124842 | 681,275 | NA | https://gwas.mrcieu.ac.uk/datasets/ieu-b-40/ |

| | | | | | | | |
|---|---|---|---|---|---|---|---|
| Coronary Artery Disease | CAD | Binary | Aragam et al. (2022) (34) | 36474045 | 714,344 | 0.04 | https://www.ebi.ac.uk/gwas/studies/GCST90132315 |
| C-Reactive Protein | CRP | Continuous | Said et al. (2022) (35) | 35459240 | 575,531 | NA | https://gwas.mrcieu.ac.uk/datasets/ebi-a-GCST90029070/ |
| Glycated Hemoglobin | HbA1c | Continuous | Garg et al. (2024) | 39261665 | 412,626 | NA | https://www.finngen.fi/en/access_results |
| Glycoprotein Acetyls | GlycAcetyl | Continuous | Karjalainen et al. (2024) (36) | 38448586 | 128,809 | NA | https://www.ebi.ac.uk/gwas/studies/GCST90301968 |
| Interleukin-6 | IL-6 | Continuous | Zhao et al. (2023) (37) | 37563310 | 14,824 | NA | https://www.ebi.ac.uk/gwas/studies/GCST90274815 |
| Leptin | Leptin | Continuous | Kilpelainen et al. (2016) (38) | 26833098 | 32,161 | NA | https://opengwas.io/datasets/ieu-a-1003 |
| Metabolic Syndrome | MetSyndr | Continuous | Park et al. (2024) (39) | 39349817 | 1,384,348 | NA | https://www.ebi.ac.uk/gwas/studi |

| | | | | | | | |
|---|---|---|---|---|---|---|---|
| | | | | | | | es/GCST90444487 |
| Stroke (all types | Stroke | Binary | Mishra et al. (2022) (40) | 36180795 | 278,025 | 0.015 | https://www.ebi.ac.uk/gwas/studies/GCST90104539 |
| Triglycerides | Triglyc | Continuous | Graham et al. (2021) (41) | 34887591 | 1,320,016 | NA | http://csg.sph.umich.edu/willer/public/glgc-lipids2021/results/ancestry_specific/ |
| Type-2 diabetes | DiabetesII | Binary | Mahajan et al. (2022) | 35551307 | 251,409 | 0.075 | https://diagram-consortium.org/downloads.html |

#### S4.2 Considerations for the Use of HDL vs LDSC

Recognizing the strengths and limitations of different approaches for estimating genetic correlations, we employed two complementary methods, namely HDL and LDSC, to maximize interpretability and cross-validate our findings. Consistent with prior observations, HDL provided more precise estimates (substantially smaller standard errors) (15). This is because HDL extends the standard LDSC framework by modelling covariances in Z statistics across multiple SNPs using a full matrix of cross-SNP LD scores. By incorporating this higher-dimensional LD structure, HDL achieves greater precision and statistical efficiency. However, despite its promise, HDL could not be applied to several closely related traits (42), likely due to collinearity issues. In contrast, LDSC yielded less precise estimates but was robust and applicable across all examined traits. This combined approach allowed us to thoroughly examine association patterns for each factor, rather than limiting our interpretation solely to statistically significant results.

### References

1. Bot M, Middeldorp CM, Geus EJC de, Lau HM, Sinke M, Nieuwenhuizen B van, *et al.* (2017): Validity of LIDAS (LIfetime Depression Assessment Self-report): a self-report online assessment of lifetime major depressive disorder. *Psychol Med* 47: 279–289.

2. Gigantesco A, Morosini P (2008): Development, reliability and factor analysis of a self-administered questionnaire which originates from the World Health Organization's Composite International Diagnostic Interview – Short Form (CIDI-SF) for assessing mental disorders. *Clin Pract Epidemiol Ment Health* 4: 8.

3. Sheehan DV, Lecrubier Y, Sheehan KH, Amorim P, Janavs J, Weiller E, *et al.* (1998): The Mini-International Neuropsychiatric Interview (M.I.N.I.): the development and validation of a structured diagnostic psychiatric interview for DSM-IV and ICD-10. *J Clin Psychiatry* 59 Suppl 20: 22-33;quiz 34-57.

4. BECK AT (1961): An Inventory for Measuring Depression. *Arch Gen Psychiatry* 4: 561.

5. Huider F, Milaneschi Y, Pool R, Maciel B de APC, Gordon SD, Rietman ML, *et al.* (2025): Genetics of Major Depressive Disorder in a Homogeneous Population with Uniform Phenotyping. https://doi.org/10.1101/2025.05.14.25325937

6. Bulik-Sullivan BK, Loh P-R, Finucane HK, Ripke S, Yang J, Patterson N, *et al.* (2015): LD Score regression distinguishes confounding from polygenicity in genome-wide association studies. *Nat Genet* 47: 291–295.

7. Devlin B, Roeder K (1999): Genomic control for association studies. *Biometrics* 55: 997–1004.

8. Bakker PIW de, Ferreira MAR, Jia X, Neale BM, Raychaudhuri S, Voight BF (2008): Practical aspects of imputation-driven meta-analysis of genome-wide association studies. *Hum Mol Genet* 17: R122-8.

9. GEORGIOPOULOS G, EVANGELOU E (2016): Power considerations for $\lambda$ inflation factor in meta-analyses of genome-wide association studies. *Genet Res* 98: e9.

10. Zaykin DV, Kozbur DO (2010): P-value based analysis for shared controls design in genome-wide association studies. *Genet Epidemiol* 34: 725–738.

11. Aguet F, Anand S, Ardlie KG, Gabriel S, Getz GA, Graubert A, *et al.* (2020): The GTEx Consortium atlas of genetic regulatory effects across human tissues. *Science* 369: 1318–1330.

12. Morey R, Zheng Y, Sun D, Garrett M, Gasperi M, Maihofer A, *et al.* (2023, October): Genomic Structural Equation Modeling Reveals Latent Phenotypes in the Human Cortex with Distinct Genetic Architecture. https://doi.org/10.21203/rs.3.rs-3253035/v1

13. Foote IF, Jacobs BM, Mathlin G, Watson CJ, Bothongo PL, Waters S, *et al.* (2022): The shared genetic architecture of modifiable risk for Alzheimer's disease: a genomic structural equation modelling study. *Neurobiol Aging* 117: 222–235.

14. Pasman JA, Allegrini AG, Tunez A, Abdellaoui A, Smit DJA, Nivard MG, *et al.* (2025): A Systematic Investigation of the Common Genetic Architecture of Substance Use Traits and the Relationship with Mental Health. *Eur Addict Res* 31: 179–196.

15. Ning Z, Pawitan Y, Shen X (2020): High-definition likelihood inference of genetic correlations across human complex traits. *Nat Genet* 52: 859–864.

16. Consortium IH 3, Altshuler DM, Gibbs RA, Peltonen L, Altshuler DM, Gibbs RA, *et al.* (2010): Integrating common and rare genetic variation in diverse human populations. *Nature* 467: 52–8.

17. Bycroft C, Freeman C, Petkova D, Band G, Elliott LT, Sharp K, *et al.* (2018): The UK Biobank resource with deep phenotyping and genomic data. *Nature* 562: 203–209.

18. Have M ten, Tuithof M, Dorsselaer S van, Schouten F, Luik AI, Graaf R de (2023): Prevalence and trends of common mental disorders from 2007-2009 to 2019-2022: results from the Netherlands Mental Health Survey and Incidence Studies (NEMESIS), including comparison of prevalence rates before vs. during the COVID-19 pandemic. *World Psychiatry* 22: 275–285.

19. Rohe K, Zeng M (2023): Vintage factor analysis with Varimax performs statistical inference. *J R Stat Soc Ser B Stat Methodol* 85: 1037–1060.

20. Huang L, Tang S, Rietkerk J, Appadurai V, Krebs MD, Schork AJ, *et al.* (2024): Polygenic Analyses Show Important Differences Between Major Depressive Disorder Symptoms Measured Using Various Instruments. *Biol Psychiatry* 95: 1110–1121.

21. Grotzinger AD, Mallard TT, Liu Z, Seidlitz J, Ge T, Smoller JW (2023): Multivariate genomic architecture of cortical thickness and surface area at multiple levels of analysis. *Nat Commun* 14: 946.

22. Baselmans BML, Weijer MP van de, Abdellaoui A, Vink JM, Hottenga JJ, Willemsen G, *et al.* (2019): A Genetic Investigation of the Well-Being Spectrum. *Behav Genet* 49: 286–297.

23. Mallard TT, Linnér RK, Grotzinger AD, Sanchez-Roige S, Seidlitz J, Okbay A, *et al.* (2022): Multivariate GWAS of psychiatric disorders and their cardinal symptoms reveal two dimensions of cross-cutting genetic liabilities. *Cell Genomics* 2: 100140.

24. Horn JL (1965): A Rationale and Test for the Number of Factors in Factor Analysis. *Psychometrika* 30: 179–185.

25. Kaiser HF (1960): The Application of Electronic Computers to Factor Analysis. *Educ Psychol Meas* 20: 141–151.

26. Gilchrist L, Spargo TP, Green RE, Coleman JRI, Howard DM, Thorp JG, *et al.* (2025): Depression symptom-specific genetic associations in clinically diagnosed and proxy case Alzheimer's disease. *Nat Ment Health* 3: 212–228.

27. Adams MJ, Thorp JG, Jermy BS, Kwong ASF, Kõiv K, Grotzinger AD, *et al.* (2024): Genome-wide meta-analysis of ascertainment and symptom structures of major depression in case-enriched and community cohorts. *Psychol Med* 54: 3459–3468.

28. Demontis D, Walters GB, Athanasiadis G, Walters R, Therrien K, Nielsen TT, *et al.* (2023): Genome-wide analyses of ADHD identify 27 risk loci, refine the genetic architecture and implicate several cognitive domains. *Nat Genet* 55: 198–208.

29. Grove J, Ripke S, Als TD, Mattheisen M, Walters RK, Won H, *et al.* (2019): Identification of common genetic risk variants for autism spectrum disorder. *Nat Genet* 51: 431–444.

30. O'Connell KS, Koromina M, Veen T van der, Boltz T, David FS, Yang JMK, *et al.* (2025): Genomics yields biological and phenotypic insights into bipolar disorder. *Nature* 639: 968–975.

31. Adams MJ, Streit F, Meng X, Awasthi S, Adey BN, Choi KW, *et al.* (2025): Trans-ancestry genome-wide study of depression identifies 697 associations implicating cell types and pharmacotherapies. *Cell* 188: 640-652.e9.

32. Trubetskoy V, Pardiñas AF, Qi T, Panagiotaropoulou G, Awasthi S, Bigdeli TB, *et al.* (2022): Mapping genomic loci implicates genes and synaptic biology in schizophrenia. *Nature* 604: 502–508.

33. Yengo L, Sidorenko J, Kemper KE, Zheng Z, Wood AR, Weedon MN, *et al.* (2018): Meta-analysis of genome-wide association studies for height and body mass index in ∼700000 individuals of European ancestry. *Hum Mol Genet* 27: 3641–3649.

34. Aragam KG, Jiang T, Goel A, Kanoni S, Wolford BN, Atri DS, *et al.* (2022): Discovery and systematic characterization of risk variants and genes for coronary artery disease in over a million participants. *Nat Genet* 54: 1803–1815.

35. Said S, Pazoki R, Karhunen V, Võsa U, Ligthart S, Bodinier B, *et al.* (2022): Genetic analysis of over half a million people characterises C-reactive protein loci. *Nat Commun* 13: 2198.

36. Karjalainen MK, Karthikeyan S, Oliver-Williams C, Sliz E, Allara E, Fung WT, *et al.* (2024): Genome-wide characterization of circulating metabolic biomarkers. *Nature* 628: 130–138.

37. Zhao JH, Stacey D, Eriksson N, Macdonald-Dunlop E, Hedman ÅK, Kalnapenkis A, *et al.* (2023): Genetics of circulating inflammatory proteins identifies drivers of immune-mediated disease risk and therapeutic targets. *Nat Immunol* 24: 1540–1551.

38. Kilpeläinen TO, Carli JFM, Skowronski AA, Sun Q, Kriebel J, Feitosa MF, *et al.* (2016): Genome-wide meta-analysis uncovers novel loci influencing circulating leptin levels. *Nat Commun* 7: 10494.

39. Park S, Kim S, Kim B, Kim DS, Kim J, Ahn Y, *et al.* (2024): Multivariate genomic analysis of 5 million people elucidates the genetic architecture of shared components of the metabolic syndrome. *Nat Genet* 56: 2380–2391.

40. Mishra A, Malik R, Hachiya T, Jürgenson T, Namba S, Posner DC, *et al.* (2022): Stroke genetics informs drug discovery and risk prediction across ancestries. *Nature* 611: 115–123.

41. Graham SE, Clarke SL, Wu K-HH, Kanoni S, Zajac GJM, Ramdas S, *et al.* (2021): The power of genetic diversity in genome-wide association studies of lipids. *Nature* 600: 675–679.

42. Zhang Y, Cheng Y, Jiang W, Ye Y, Lu Q, Zhao H (2021): Comparison of methods for estimating genetic correlation between complex traits using GWAS summary statistics. *Brief Bioinform* 22: bbaa442.
